# Decoding pan-cancer treatment outcomes using multimodal real-world data and explainable artificial intelligence

**DOI:** 10.1101/2023.10.12.23296873

**Authors:** Julius Keyl, Philipp Keyl, Grégoire Montavon, René Hosch, Alexander Brehmer, Liliana Mochmann, Philipp Jurmeister, Gabriel Dernbach, Moon Kim, Sven Koitka, Sebastian Bauer, Nikolaos Bechrakis, Michael Forsting, Dagmar Führer-Sakel, Martin Glas, Viktor Grünwald, Boris Hadaschik, Johannes Haubold, Ken Herrmann, Stefan Kasper, Rainer Kimmig, Stephan Lang, Tienush Rassaf, Alexander Roesch, Dirk Schadendorf, Jens T. Siveke, Martin Stuschke, Ulrich Sure, Matthias Totzeck, Anja Welt, Marcel Wiesweg, Hideo A. Baba, Felix Nensa, Jan Egger, Klaus-Robert Müller, Martin Schuler, Frederick Klauschen, Jens Kleesiek

**Author notes:** Contributed equally (Julius Keyl, Philipp Keyl, Klaus-Robert Müller, Martin Schuler, Frederick Klauschen, Jens Kleesiek).

## Abstract

Despite advances in precision oncology, clinical decision-making still relies on limited parameters and expert knowledge. To address this limitation, we combined multimodal real-world data and explainable artificial intelligence (xAI) to introduce novel AI-derived (AID) markers for clinical decision support.

We used deep learning to model the outcome of 15,726 patients across 38 solid cancer entities based on 350 markers, including clinical records, image-derived body compositions, and mutational tumor profiles. xAI determined the prognostic contribution of each clinical marker at the patient level and identified 114 key markers that accounted for 90% of the neural network’s decision process. Moreover, xAI enabled us to uncover 1,373 prognostic interactions between markers. Our approach was validated in an independent cohort of 3,288 lung cancer patients from a US nationwide electronic health record-derived database.

These results show the potential of xAI to transform the assessment of clinical parameters and enable personalized, data-driven cancer care.

## Introduction

Despite the vast amount of multimodal clinical data currently available for each patient in modern healthcare, the promise of personalized medicine has yet to be realized. Every year, numerous studies are published on individual prognostic markers in oncology. However, most of these single-marker studies do not provide sufficient insight into the complex interplay of patient- and tumor-specific variables that determine a patient’s prognosis.^1^ As a result, many of the proposed tools are not used in clinical practice or do not consider the patient’s entire clinical data reflecting the unique disease context.^2,3^ A promising strategy to overcome this limitation is to integrate clinical data from multiple sources, such as medical history, laboratory test results, imaging data, and omics analyses.^1,4^ Advances in machine learning and the increasing availability of digitally accessible data made it possible to model complex relationships between prognostic markers on a large scale.^1,5–9^ Together with recent methods for understanding the decision-making of such models, referred to as explainable artificial intelligence (xAI), this makes it possible to assess individual patient prognosis and unravel the contribution of each parameter.^10–14^

In this study, we leveraged these advances by proposing an approach for decoding prognostic hallmarks based on large-scale real-world data. We modeled patient outcomes using a deep neural network and applied the xAI method layer-wise relevance propagation (LRP) to disentangle how each piece of clinical information contributed to an individual patient’s prognosis.^5,12^ Our dataset comprises multimodal data from 15,726 patients across 38 cancer entities undergoing systemic treatment. The data include clinical examination, laboratory tests, clinical records, computed tomography imaging (CT)-derived body composition, and genetic data. Until now, many existing clinical predictors have been cancer-entity specific and not designed to incorporate cross-cancer associations. However, genetic findings from pan-cancer studies are challenging this strict classification. Available data suggest that similarities between patients extend beyond the histological tumor type, leading to an increasing number of basket trials that include patients with different cancer entities.^15–20^

Training our deep-learning approach on a pan-cancer dataset enabled the neural network to learn prognostic relationships that extend across cancer entities. This facilitates the development of a comprehensive model that reveals clinically relevant biomarker signatures without any prior knowledge. As a result, our approach can aid clinicians in prioritizing critical patient-specific information and optimizing therapeutic strategies. At the cohort level, the modularity of our approach allowed us to assess how the importance of each clinical marker varied in different disease contexts and between arbitrary patient groups. A striking application is the development of explainable Kaplan-Meier (xKM) plots that illustrate the evolution of marker importance during disease progression. We confirmed the reproducibility and validity of this xAI approach on an external real-world dataset comprising 3288 lung cancer patients from a US nationwide, electronic health record-derived, de-identified database.

The growing abundance and accessibility of real-world data is increasingly revealing its potential for clinical application, paving the way for more precise and personalized treatments. In this study, we move further and demonstrate the ability of xAI to decode patient outcomes and provide tailored treatment guidance based on multimodal real-world data.

## Results

### Cohort definition

We retrospectively evaluated data from 150,079 cancer patients with available medical records treated at the West German Cancer Center of the University Hospital Essen, one of Germany’s largest academic comprehensive cancer centers. Of these, 15,726 patients who received systemic cancer treatment between 1 April 2007 and 22 July 2022 (median: November 2016) were included in the final analysis (**Suppl. Fig. 1**). The most frequent cancer entities were lung cancer (n=4,320), sarcoma (n=1,578), and breast cancer (n=1,223; for details see **Suppl. Table 1)**. Metastatic status (M status) was available in a structured format at baseline for 7,965 patients. Of those, 5,606 patients were treated for metastatic disease (M1), and 2,359 patients received systemic therapy for localized or locally advanced cancers (M0). In 5,395 patients, body composition was automatically assessed from abdominal CT images taken before treatment initiation.^21,22^ In total, we included 350 parameters in our analysis, consisting of different modalities and both patient- and tumor-specific parameters (see methods for a detailed description of the parameters).

### Models trained on multimodal pan-cancer data accurately predict survival and treatment outcomes

Two neural networks were trained to predict overall survival (OS) and time-to-next-treatment (TTNT) for each patient based on their medical profile at the time of first in-house systemic treatment. We demonstrated the reliability of the neural networks by performing a five-fold cross-validation. For each fold, two neural networks were trained (80% of samples), validated (10% of samples), and tested (10% of samples) for the prediction of OS and TTNT, respectively.

The survival model achieved an average concordance index (C-index) on the pan-cancer dataset of 0.762 (range across folds: 0.758-0.764) for OS prediction and 0.711 (range: 0.702- 0.718) for TTNT prediction across all cancer entities (**Fig. 2A**). When the model performance was tested independently for each cancer entity with at least 20 patients in each fold’s test set, the predictive performance varied. For OS, the highest C-index was achieved for ocular cancers (0.804, range: 0.771-0.860), while the highest C-index prediction of TTNT was achieved for rectal cancers (0.756, range: 0.644-0.800).

**Figure 1:**
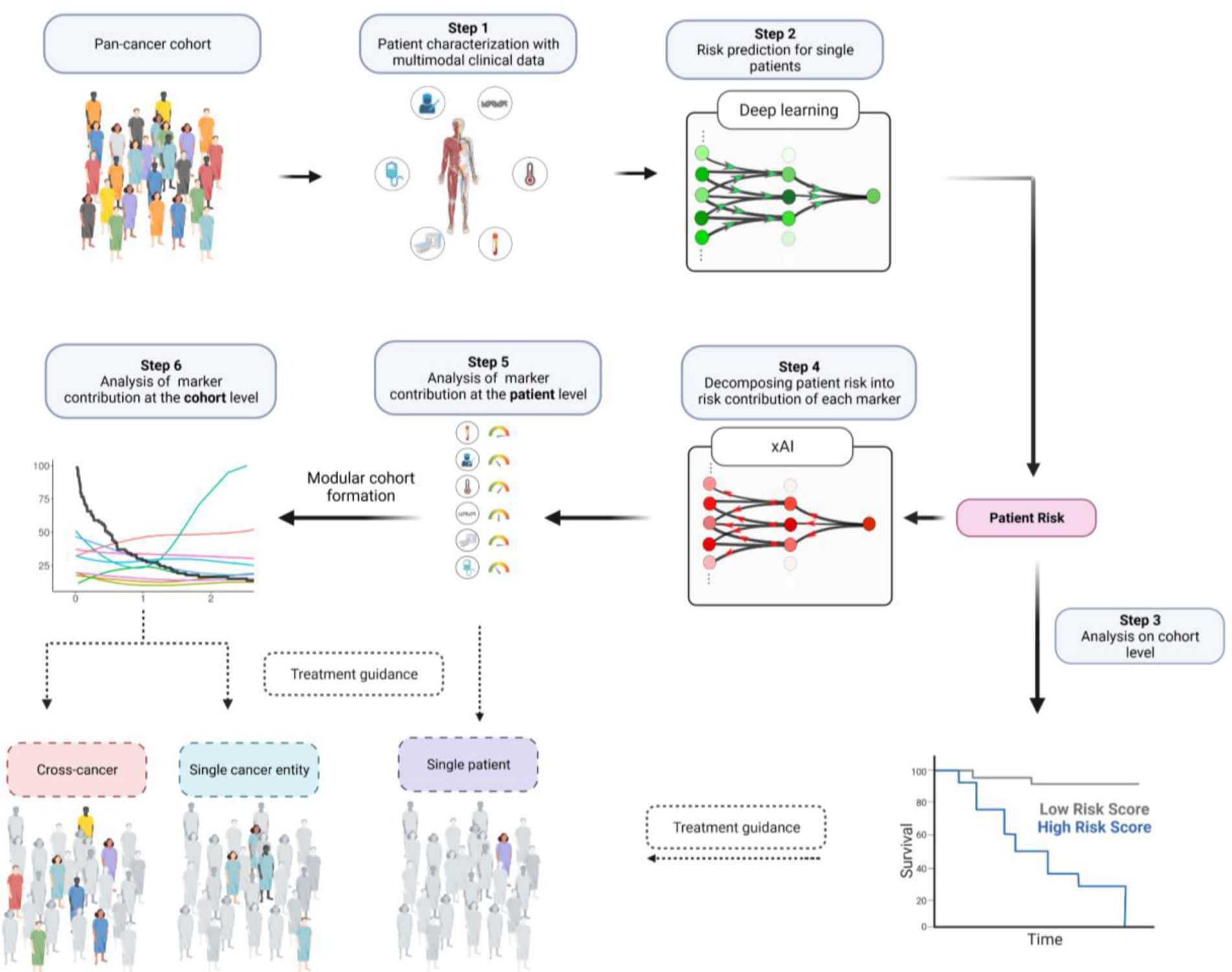
Overview of the xAI-based workflow for decoding treatment outcomes at the patient and cohort level. Following the collection of multimodal pan-cancer data, each patient’s risk score is predicted by deep learning and enables patient stratification. xAI then decomposes the patient risk into the individual contributions of each marker. This enables treatment guidance at the patient and cohort level.

**Figure 2:**
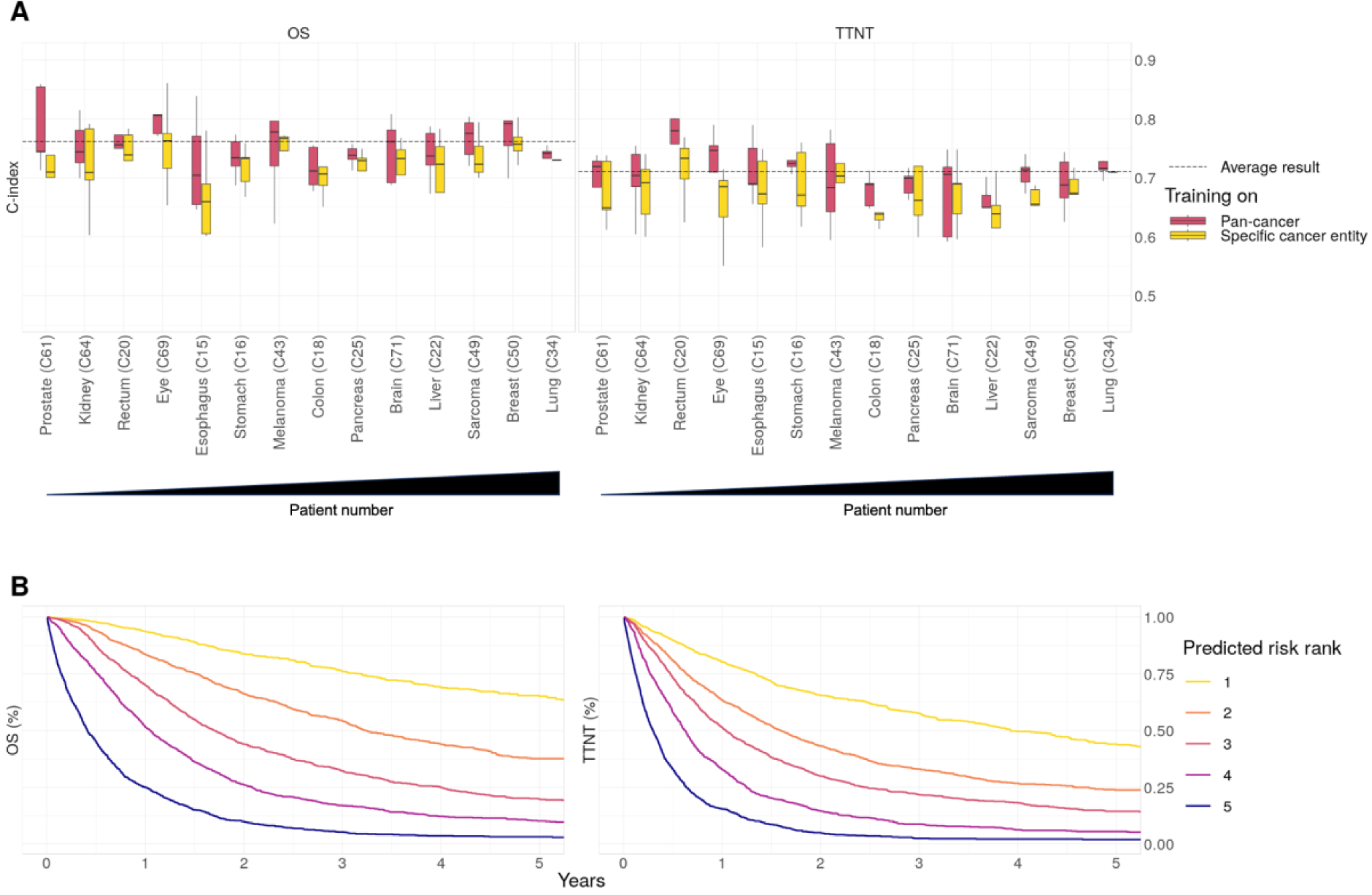
Prediction of prognosis following training on pan-cancer real-world data. **A:** Concordance index for predicting overall survival (OS) and time-to-next-treatment (TTNT) in five-fold cross- validation. The dashed line indicates the prediction result over all cancer entities and folds. Box plots show prediction results for individual cancer entities with at least 20 patients in the test set of each fold after training the neural network on all cancer entities (red) or the specific cancer entity (yellow). Cancer entities are ordered from left to right by ascending patient numbers. **B:** Kaplan-Meier plots for overall survival and time-to-next-treatment in the pan-cancer dataset for patients of the combined test sets (7,861) patients. Patients were stratified into five risk groups according to the risk predicted by the neural network.

Training models on the pan-cancer dataset, as opposed to exclusively training on single cancer entities, significantly improved model performance for both OS (mean C-index: 0.75 vs. 0.72, p<0.001) and TTNT (mean C-index: 0.70 vs. 0.68, p<0.001). Only in melanoma patients, the mean results (mean C-index for OS: 0.74 vs. 0.75, mean C-index for TTNT: 0.69 vs. 0.7, p-value>0.05) were better when the training was performed on the melanoma cohort compared to training on the pan-cancer cohort. The advantage of the pan-cancer model over the single-entity models was particularly striking for cancer entities with low patient numbers (**Fig. 2A**). This suggests that the pan-cancer model used prognostic information shared by different cancer entities to provide meaningful predictions even for patients with rare cancers.

After training on a large and granular real-world pan-cancer dataset, both neural networks for predicting OS and TTNT were able to stratify patients from the test sets into distinct cross-cancer risk groups (**Fig. 2B**).

### xAI reveals the complex relationships between markers and prognosis

After developing reliable outcome prediction models, we applied xAI to unravel how clinical information of individual patients influences the neural networks in assessing prognosis. We chose the xAI method layer-wise relevance propagation (LRP) because it allows for the computation of robust explanations at low computational cost for individual patients.^12^ LRP computed for each patient the risk contribution (RC) of every clinical parameter, such as laboratory markers or comorbidities, to the predicted favorable or unfavorable outcome. This results in ‘AI-derived’ (AID) markers with two dimensions, the original marker value and its LRP-assigned RC. A positive RC indicates a contribution to an adverse outcome and a negative RC indicates a contribution to a favorable outcome.

By analyzing the AID markers across all patients, it was possible to investigate how the neural network evaluated the relationship between the marker and its contribution to the patient’s risk (**Fig. 3A**). For example, increasing age and elevated CRP strongly contributed to predicting an unfavorable prognosis. In contrast, high fT3, high PD-L1 TPS, and higher CT-derived abdominal muscle volume contributed to predicting a favorable prognosis.

**Figure 3:**
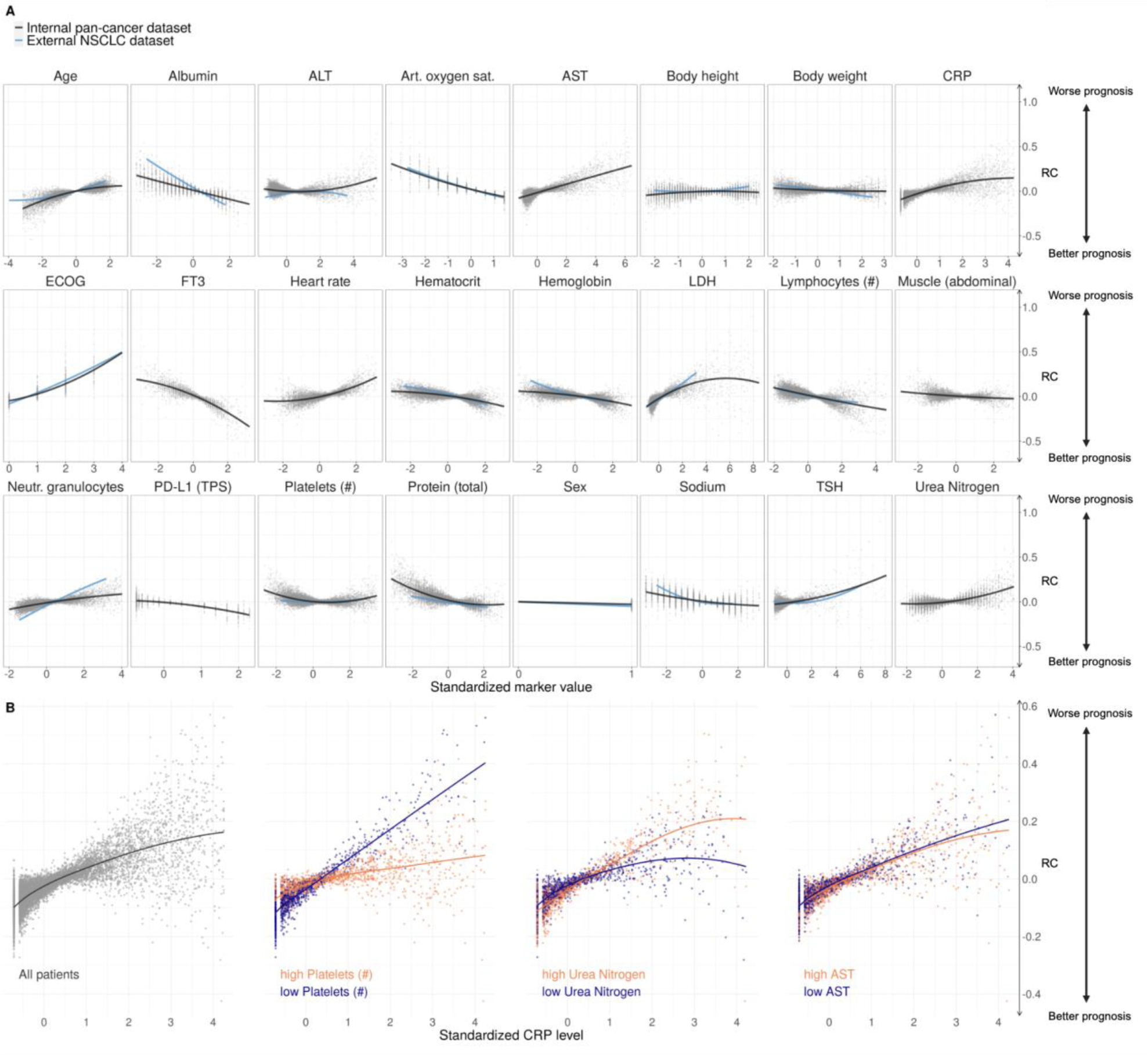
Contribution of clinical markers to the prediction of OS. A: Marker risk contribution (RC) on the OS prediction. Each point represents one marker value for one patient versus the LRP-assigned RC (y axis) to the patient’s prognosis. Marker values are standardized. B: The risk contribution of CRP depended on the value of other markers. The left plot shows the standardized CRP level and LRP-assigned RC for all patients. The right three plots depict the patients for whom the three selected markers platelet count, urea nitrogen, and AST were in the highest or lowest 10% quantile.

We validated the results for a subset of markers using external data from 3288 patients with non-small cell lung cancer (NSCLC) provided by Flatiron Health. Upon applying our approach to the external dataset, we found a strong correlation between the linearized slopes of RCs on the internal and external datasets (Pearson’s r=0.9, p<0.001, **Suppl. Fig. 2**). Thus, xAI predicted a comparable impact of markers on patient risk in both datasets. To confirm if the fundamental results of LRP matched conventional models, we examined the simplified linearized effect predicted by xAI against a standard Cox Proportional Hazards model. Our analysis revealed that the relationships computed on the internal and external datasets strongly correlated to the hazard ratios of each marker (internal dataset: Pearson’s r=0.93, p<0.001, external dataset: Pearson’s r=0.97, p <0.001, **Suppl. Fig. 3**).

Notably, the RC of a marker varied widely even when different patients had the same marker value. By utilizing LRP, it becomes possible to explain some of the variance in RC by marker interactions (**Fig. 3B**). We observed how the RC of CRP varied depending on the values of additional ‘secondary’ parameters. Out of 8,294 examined marker pairs, 1,373 (16.6 %) showed significant interactions according to a mixed-effects model. For example, high CRP levels were assigned a high RC, particularly when platelet counts were low (Δ RC slopes: - 0.07, p<0.001). CRP had less influence on the predicted risk when the platelet count was high. While the prognostic significance of elevated CRP levels and platelet counts is known, the exact interaction has not yet been described.^23^ The impact of blood urea nitrogen (BUN) on the RC of CRP was less pronounced (Δ RC slopes: 0.03, p<0.001). Here, a higher CRP level was associated with a particularly high RC in patients with high BUN levels. In contrast, the RC of CRP was independent of aspartate aminotransferase (AST) (Δ RC slopes: -0.006, p=1.0). For results on TTNT, see **Suppl. Fig. 4**.

The statistically significant interactions between the features present in the internal and external datasets showed a high level of similarity in the external dataset (Pearson’s r=0.59, p=0.021. **Suppl. Fig. 5**). To confirm that the fundamental interaction results observed with xAI were consistent with conventional models, we examined the simplified linearized effect over the LRP-assigned RC against a mixed-effects Cox Proportional Hazards model.

Here, the direction of interactions derived from xAI matched the interactions observed with the Cox regression models in the internal and external datasets (r=0.91, p=0.03 and r=0.69, p=0.009, **Suppl. Fig. 6**). Based on these results, we concluded that the LRP approach was highly reproducible across various datasets as well as consistent with established statistical models that simplify relationships. However, the xAI approach’s full potential extends beyond this and enables nonlinear RC assignments for individual patients, taking into account their unique disease context.

### AI-derived (AID) markers for patient-level treatment guidance

AID markers, the combination of a marker value with its LRP-assigned risk contribution, enhance the clinical information available to healthcare professionals by incorporating the contextual risk associated with each marker. A ‘clinician’s guide’ can clearly present the AID marker profile of individual patients.

In Fig. 4, we show representative results that illustrate a potential real-world use case of the ‘clinician’s guide’ for four different patients. In patient 1, age, BMI, body weight, and fT3 values contributed unfavorably to the overall prognosis, while the high lymphocyte and platelet counts were assigned a favorable (negative) RC. The patient’s prognosis deteriorated with impaired breathing, aphagia, pain, and an advanced T and M stage. Among the different distant metastases, liver metastases were identified as particularly unfavorable compared to lung and bone metastases. Overall, the neural network therefore predicted a highly adverse outcome for this patient based on all available data. In patient 2, lymphocytopenia and older age particularly contributed to a poor prognosis. However, this patient had few comorbidities, with pleural effusion having the strongest unfavorable impact. The absence of liver metastases and the treatment with pembrolizumab were assigned a favorable RC, and the overall risk was considered intermediate. Notably, patient 3 had elevated CRP levels, which is conventionally associated with a potentially dangerous patient condition requiring increased monitoring. However, xAI does not consider this parameter to be detrimental in this particular case. As shown before, this may be due to this patient’s high platelet and low urea nitrogen levels. Patient 4 showed medium visceral adipose tissue (VAT), contributing favorably, and low subcutaneous adipose tissue (SAT), contributing adversely. With few comorbidities and no metastases, the overall prognosis was favorable.

**Figure 4:**
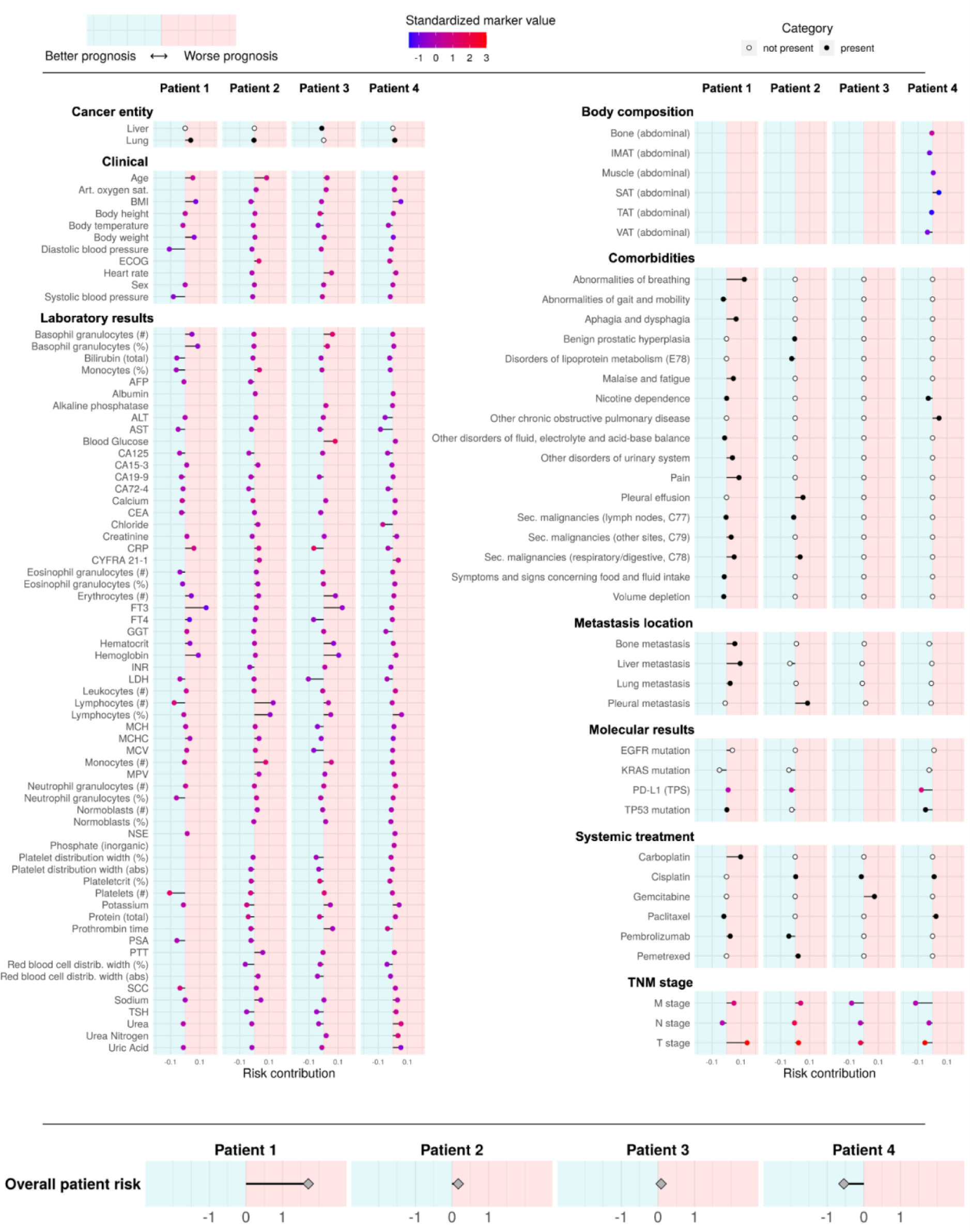
Clinician’s guide showing the contribution of each marker to overall risk at the patient level. Representative results of four patients are presented. The x axis indicates the marker’s risk contribution towards higher (right/positive) or lower (left/negative) risk. Colors indicate the presence (black) or absence (white) of cancer entities, comorbidities, metastasis locations, and systemic treatment. For markers with ordinal or continuous scales, the point color indicates the marker value for the respective patient. For continuous markers, marker values are standardized. The predicted overall patient risk is displayed at the bottom. Body composition markers: Abdominal volumes of visceral adipose tissue (VAT), total adipose tissue (TAT), subcutaneous adipose tissue (SAT), intermuscular adipose tissue (IMAT), muscle, bone.

### Evaluation of established scoring systems

Our results illustrated the limitations of single marker-based outcome prediction and emphasized the importance of prognostic parameters to be considered in the disease context characterized by other markers. In clinical routine, however, it is common to rely on a few scoring systems, such as the TNM stage, to assess prognosis and guide treatment. Based on these scoring systems, patients are usually rigidly categorized, regardless of fundamental differences such as sex, nutritional status, or comorbidities.

To evaluate the dependency of a score on this disease context, we analyzed the correlation between the score and the LRP-assigned RC (**Suppl. Fig. 7**). For Eastern Cooperative Oncology Group performance status (ECOG PS) (r=0.87), M stage (r=0.92), and N stage (r=0.76), higher scores correlated with higher computed RC on average, indicating a consistent influence on the prognosis independent of other markers. The weak correlation of tumor grade (r=0.02) and T stage (r=0.07) with their RC suggested that they should be interpreted in the context of additional markers.

### Assessment of marker importance at the cohort level

In a multimodal real-world dataset reflecting clinical care, there are expected to be both sideline markers of low prognostic relevance and critical markers that are highly relevant across patients. To measure the marker importance (MI) in a cohort, we calculated the absolute value of the RC in consistency with other methods in the field.^13^ We found that 90% of LRP scores were assigned to the 114 most important markers out of 350 (**Suppl. Fig. 8A**). Across all patients, the most important markers for the prediction of OS were C-reactive protein level (CRP, mean MI: 0.071), free triiodothyronine (fT3, mean MI: 0.066), ECOG PS performance status (mean MI: 0.061), M stage (mean MI: 0.058) and LDH (mean MI: 0.055, see **Suppl. Fig. 9A**). These results are consistent with previously reported findings.^24–27^

Estimating the contribution of a particular comorbidity or intervention to a patient’s overall prognosis is difficult as each is a rare event. LRP can assess the influence of comorbidities, defined by ICD codes, and medical interventions, defined by the German operation and procedure classification system (OPS), in the disease context (**Suppl. Fig. 10A)**. Due to the scarcity of each comorbidity, MI was not informative here, which is why we report the mean RC of affected patients. The comorbidities that contributed the most to the prediction of a poor outcome were pain (mean RC: 0.064), respiratory abnormalities (mean RC: 0.064), ascites (mean RC: 0.056), secondary malignant neoplasm of the respiratory or digestive tract (mean RC: 0.048), and pleural effusion (mean RC: 0.046). Notably, some diagnoses contributed favorably to the overall prognosis (e.g., heart failure, gastritis, and duodenitis). The interventions that were assigned the highest RC were ureteral stenting (mean RC: 0.074), which may indicate a stenotic process, and meningeal reconstruction (RC: 0.049).

For results on TTNT, see **Suppl. Fig. 8B, 9B, and 10B**.

### Cross-cohort comparison of prognostic markers

Model training on a pan-cancer dataset and sample-wise explanations obtained by LRP allowed us to investigate how the MI of a marker differed between patient subgroups (**Fig. 5**). In several cases, we confirmed well-established relationships between markers and cancer entities.

**Figure 5:**
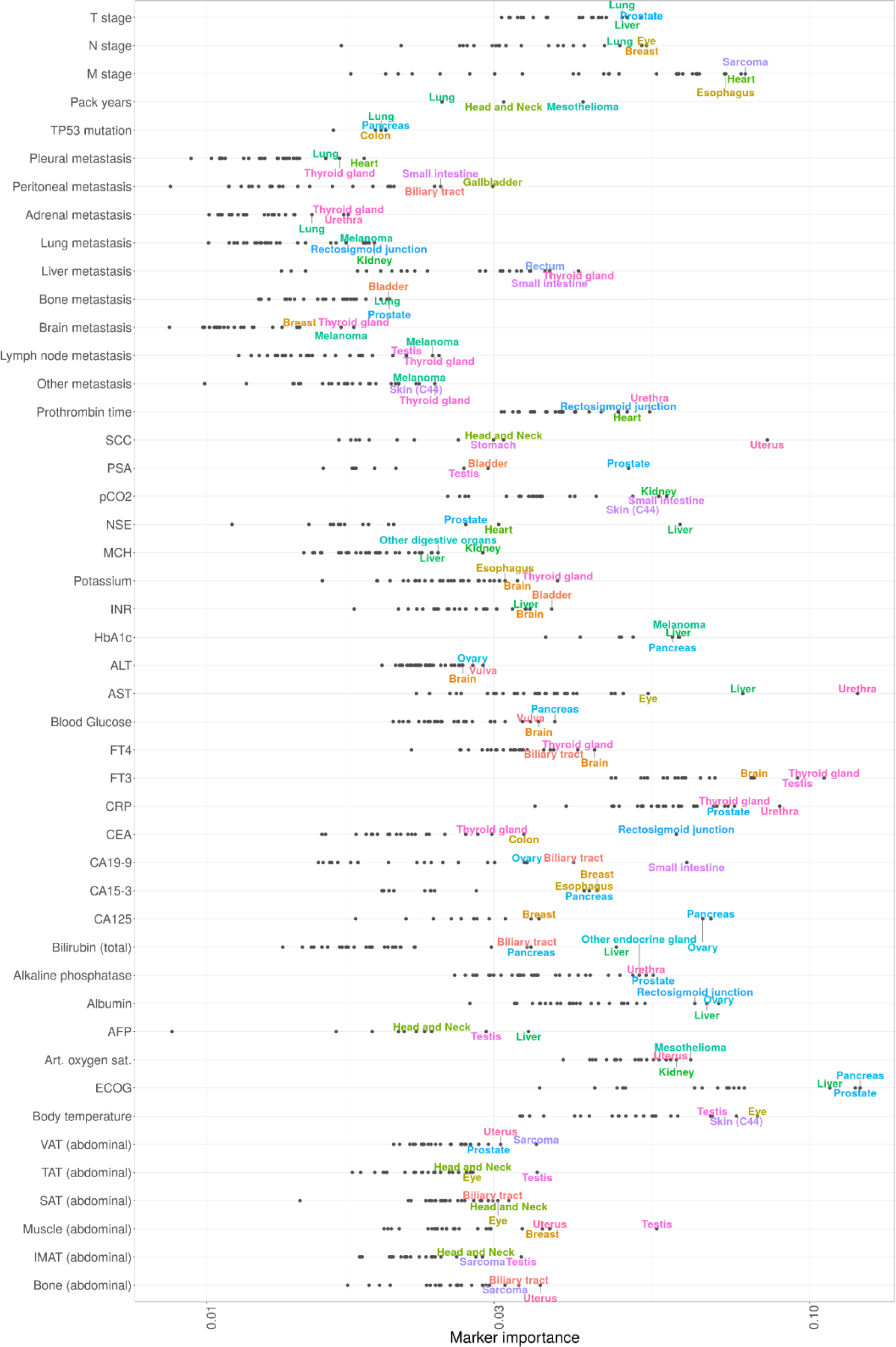
The relationship between mean marker importance (MI) of selected markers and cancer entities. The x axis shows the MI on a logarithmic scale. The three cancer entities with the highest marker MI are annotated for each marker. Body composition markers: Abdominal volumes of visceral adipose tissue (VAT), total adipose tissue (TAT), subcutaneous adipose tissue (SAT), intermuscular adipose tissue (IMAT), muscle, bone. ECOG PS had the highest MI in pancreatic, prostate, and liver cancers, whereas fT3 was particularly important for thyroid, testicular, and brain cancers.

In our results, CA19-9 had the highest MI in cancers of the small intestine, pancreas, and biliary tract.^28,29^ The presence of liver metastases was most relevant for cancers of the thyroid gland, rectosigmoid junction, and additional digestive tract cancers.^30,31^ Bilirubin emerged as an essential marker for liver, pancreatic, or biliary tract cancers, while fT3 and fT4 were most important in thyroid, testicular, and brain cancers.^32–34^ Prostate-specific antigen (PSA) had the highest MI in prostate cancer, followed by the locally adjacent entities bladder and testicular cancer. Abdominal muscle volume, as determined by CT-based body composition analysis, was most impactful in vulvar, uterine, and testicular cancers. HbA1c was most important in cancers of the pancreas, the liver, and in melanoma. The tumor marker CEA had the highest MI in cancers of the rectosigmoid junction, the colon, and the thyroid, while the ECOG PS was particularly important for pancreatic, prostate, and liver cancers. Interestingly, AST had very high MI for urethral cancer, followed by liver and ocular cancer (mainly uveal melanoma).

For results on TTNT, see **Suppl. Fig. 11**.

### Explainable Kaplan-Meier plots (xKM) visualize the evolution of marker importance during disease progression

Having examined the cancer entity-specific impact of markers on prognosis, we further explored their varying importance for prognostication during disease progression. Ordering the deceased patients according to OS, we could follow the LRP-assigned marker importance along a pseudo timeline and observed distinct changes over the course of treatment (**Fig. 6**). ECOG PS, CRP, and LDH levels were highly prognostic markers throughout disease progression across all cancer entities. The prognosis of patients with a short OS was strongly influenced by total serum protein concentration, which may reflect the relevance of organ dysfunction at this stage of the disease, particularly of the liver and kidneys. The coagulation parameter prothrombin time and oxygen saturation were highly prognostic in patients with long OS but contributed much less to the prognosis of patients with short OS. M stage had an overall decisive marker importance, which decreased for disease stages with short OS.

**Figure 6:**
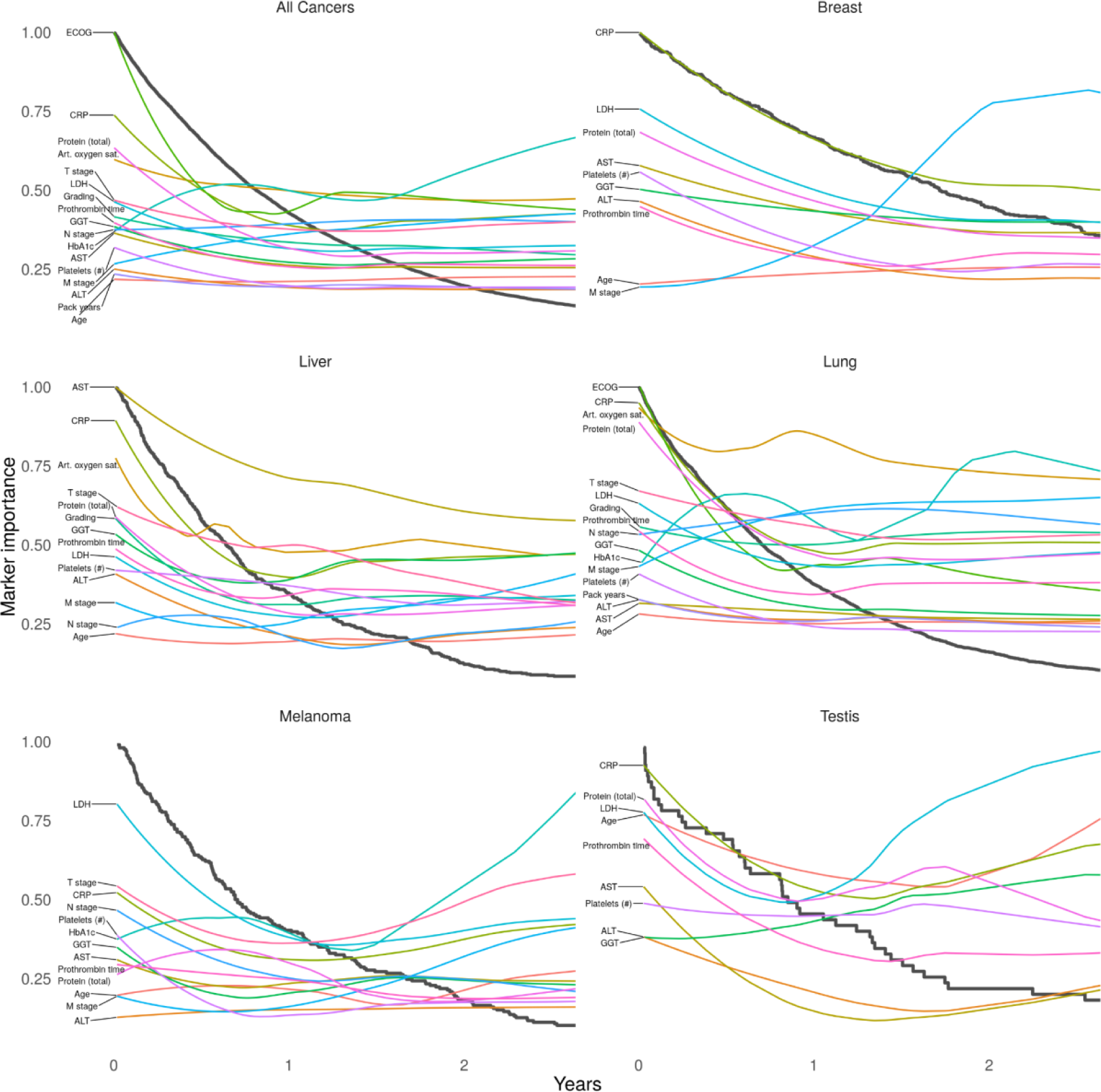
Explainable Kaplan-Meier (xKM) plots depicting the importance of diagnostic markers during disease progression. Black lines represent Kaplan-Meier plots, while the colored lines visualize the change in marker importance (MI) for patients with different survival times. MI lines are scaled between zero and one. Only deceased patients were included in this analysis.

Our modular approach allowed us to generate explainable Kaplan-Meier (xKM) plots of patient subgroups with different prognoses. We identified parameters that separated prognostic subgroups in a given cancer entity. In lung cancer, arterial oxygen saturation had the highest MI for most patients, but for patients with short survival protein expression and CRP became even more critical. Metastasis (M stage) generally had higher MI than lymph node metastasis and tumor stage. Interestingly, the importance of metastasis decreased during disease progression and was overtaken by T stage and N stage in patients who survived only a few months. LDH had exceptionally high MI in testicular cancer and melanoma, which is well known in the literature.^35,36^ The MI of the latter increased during disease progression. In the liver, the MI of AST, total protein, GGT, prothrombin time, and LDH increased during disease progression. ALT was less important for patients who survived more than one year.

Next, we examined the prognostic impact of cancer-specific biomarkers (**Suppl. Fig. 12**). PD- L1 TPS was the most important cancer-specific marker for lung cancer prognosis, which aligns with the efficacy of immune checkpoint inhibitor therapy.^37^ In head and neck cancer, the tumor marker SCC had a high marker importance that increased during disease progression. In liver cancer, the tumor marker AFP was of high MI throughout disease progression, but CA19 -9 and CA125 became more important towards the end of life.

xKM plots for OS can be viewed interactively online at **xkaplanmeier.streamlit.app**.

For results on TTNT, see **Suppl. Fig. 13** and **Suppl. Fig. 14**.

## Discussion

Personalized medicine requires a comprehensive characterization of individual patients, which cannot be achieved by conventional scoring systems based on limited sets of markers.^1,4^ Despite the extensive routine diagnostic data available for each patient, current clinical tools only include small subsets of these parameters in a limited number of cancer entities.^2,3^ Previous studies have started to show the potential of utilizing multimodal data to predict individual patient prognosis using public databases.^7,8,17^ In this study, we utilized multimodal routine clinical data from 15,726 patients with solid cancers undergoing systemic treatment at a large comprehensive cancer center to uncover the complex mechanisms that determine a patient’s prognosis. By incorporating a wide range of parameters and cancer entities, we developed powerful prognostic models for OS and TTNT prediction. We found that the models benefited from training on patients of both the same and different cancer entities, resulting in the successful stratification of patients into cross-cancer risk groups. This is consistent with the growing trend to guide treatment based on predictive biomarkers across cancer entities.^18–20^ Using xAI, our study provided a comprehensive understanding of the factors contributing to a treatment outcome. Without using any prior knowledge, xAI characterized how each patient’s prognosis was determined by their individual marker profile and identified CRP, fT3, M status, and ECOG performance status as the most important factors across all patients. While xAI methods have already demonstrated their reliability for various tasks in and outside of the medical field, we have verified our results on an independent cohort of NSCLC patients. Our results showed excellent reproducibility between datasets and were highly consistent with conventional methods.

In the medical domain, xAI has previously been applied to validate the model performance or assess feature importance across cohorts.^17,22,38^ Few studies have made use of patient-wise xAI explanations.^17^ Here, we built on xAI to contextualize complex multimodal patient data and systematically reveal the underlying mechanisms driving a patient’s disease progression. We leveraged the potential of xAI for patient-level explanations and developed AI-derived (AID) markers with dual dimensions, the original marker value and the xAI assigned risk contribution. This approach has practical implications, as it can guide clinicians in identifying patient characteristics that contribute most to adverse outcomes and therefore require special attention. The application is not limited to cancer patients and may be particularly interesting in emergencies where rapid treatment decisions are critical. By systematically comparing these AID markers among patients, we found that the marker importance varied widely across cancer entities and during disease progression, which we visualized using explainable Kaplan-Meier (xKM) plots. Our findings show that prognostic associations are not static and that different markers may be critical depending on the cancer entity and the individual disease setting. Prognostic tools based on predefined parameters and assessment criteria are therefore insufficient to capture the complex prognostic relationships and may lead to inaccurate prognostic estimates. In contrast to traditional statistical methods, xAI can build on all available data to assess the complex setting of individual patients. In clinical practice, medical data from various sources, including multi-omics data, are becoming more widely accessible for research purposes.^4,39^ xAI methods can be a viable solution to facilitate data analysis for routine clinical care and research, provided that common pitfalls are addressed.

Confounding is one of the most common challenges in retrospective real-world data analysis. We aimed to reduce confounding effects caused by correlating input features by applying dropout regularization not only to the neural network weights but also to the input.^40^ In a real-world data setting, confounding can also be introduced by documentation. For instance, gastritis or duodenitis are not expected to positively impact the patient’s prognosis. However, the documentation of these non-cancer comorbidities may have suggested the absence of an acute life-threatening condition. Also, selection bias should be considered in real-world data studies. In this proof-of-concept study, we enrolled only patients receiving systemic cancer therapy. While this cohort provides well-structured treatment data, it is more likely to include patients with advanced disease. Particular caution is also needed when interpreting the RC assigned to the different treatments, as the non-randomized selection of treatments may lead to statistical bias.

In clinical trials, randomization prevents certain forms of confounding and bias. Real-world studies combined with xAI will therefore not replace RCT but may generate new data-driven hypotheses and inform RCT design.^41^ Due to the versatility of our approach, it can be applied to virtually any dataset containing clinical parameters and endpoints in future studies. Further image, text, or omics data could be integrated seamlessly. Since it is not limited to real-world data, RCT designed for specific clinical settings could also directly integrate our xAI framework.

In summary, we demonstrate a novel xAI-based approach for large-scale multimodal data analysis of prognostic relationships in a real-world setting. Given the increasing influence of multi-omics and image data on patient management and therapy selection, xAI approaches hold great potential for precision medicine.

## Methods

### Study design

Electronic health records from 150,079 cancer patients treated at University Hospital Essen were retrospectively evaluated. Of these, we included 15,726 patients who underwent systemic cancer treatment at University Hospital Essen between 1 April 2007 and 22 July 2022 in this study. Overall survival (OS) was defined as the time from initiation of systemic treatment to death from any cause. Time to next treatment (TTNT) was defined as the time from initiation of systemic treatment until initiation of next line of systemic treatment or death from any cause. Patients for whom no date of death was available were censored at the date of the last follow-up. The study was approved by the Ethics Committee of the Medical Faculty of the University of Duisburg-Essen (No. 21-10347-BO).

### Data acquisition

All medical data were retrieved from the smart hospital information platform (SHIP) of University Hospital Essen. In SHIP, medical data is stored in FHIR format and can be collected based on specific queries. The various subsystems at Essen University Hospital, e.g., for laboratory values or electronic medication administration, automatically transfer the data to SHIP. First, all patients with solid tumors were collected based on ICD codes (C00-C75). Then, patients who received intravenous or oral cancer treatment documented in SHIP were selected. Further inclusion criteria were: Initiation of systemic therapy since 1 April 2007 and a minimum age of 18 years at the initiation of cancer treatment. A detailed overview of the patient enrollment process can be found in the supplementary material (**Suppl. Fig. 1**).

For the resulting cohort of 15,726 patients, further clinical data were retrieved from SHIP. To ensure a balance of the most recent data with the fewest missing values in our dataset, we defined different time windows for querying the parameter sets relative to the start of systemic cancer treatment. Listed below are all of the queried parameter sets used to create our dataset, along with the time windows where applicable:

● Systemic cancer treatment (first recorded in SHIP): For each patient, the substances of the first line of therapy administered in our cancer center were retrieved. The data originate from our electronic medication administration system. In total: 48 parameters.
● Demographics: Age, sex, height (in total: Three parameters)
● Body composition (max. two months before treatment): In addition to weight and BMI, we included abdominal body composition, which was automatically obtained from CT images, to accurately assess the physical condition of patients. We retrieved abdominal CT images with a maximum interval of two months before treatment initiation and used a deep learning model to automatically measure muscle, bone, and different fat volumes (subcutaneous, visceral, intermuscular, and total adipose tissue).^21^ The collected markers were divided by the number of abdominal CT slices to ensure patient comparability. In total: Eight parameters.
● Cancer entity (C0-75): For each patient, exactly one cancer entity was queried for which they were receiving treatment. In total: 60 parameters
● Prior diagnoses (any before treatment): We selected all ICD-10 codes (except C0-C75) that were present in at least 200 patients. In total: 68 parameters
● Prior medical interventions (any before treatment): We used the German operation and procedure classification system (OPS) to identify prior medical interventions. We selected all OPS codes that were present in at least 200 patients. In total: 50 parameters
● Staging (max. one year before treatment): T, N, and M status were obtained from tumor board documentation. In total: Three parameters
● Metastasis localization (any before treatment): Tissue affected by metastasis, if any. In total: Nine parameters
● Clinical parameters (max. two weeks before treatment): Oxygen saturation, body temperature, heart rate, systolic and diastolic blood pressure. In total: Five parameters
● ECOG PS (max. three months before treatment): ECOG PS was obtained from tumor board documentation. In total: One parameter
● Laboratory parameters (max. two weeks before treatment): We selected all parameters that were present in at least 20% of patients (62 parameters), plus nine others (mainly tumor markers) that we considered particularly relevant for subgroups. In total: 71 parameters
● Pathology: Cancer subtype beyond ICD-10 classification, histologic tumor grade, immunohistochemical results, and somatic tumor mutations. In total: 22 parameters
● Smoking status: Smoking status (smoker/ non-smoker) and, if available, pack-years of smoking. In total: Two parameters

The endpoints OS and TTNT were automatically extracted from SHIP.

### Data preprocessing

Outliers, defined as >3 standard deviations from the mean, were removed for continuous parameters. Continuous parameters were prestandardized to zero mean and unit variance. Categorical scores were encoded on an ordinal scale (e.g., ECOG PS as 0-4, metastasis as 0-1). Diagnoses (ICD codes), cancer entities, interventions (OPS codes), and systemic cancer treatments were one-hot encoded (0=not present, 1=present). This resulted in a total of 350 parameters for the final dataset. For further analysis and description of differences between cancers, the cancer representations were summarized into more general cancer entities (**Suppl. Table 1**). To account for missing values while simultaneously keeping the ability to explain the present clinical markers, we applied feature expansion: *x* → (*x*, 1 − *x*). Missing values were set to (0,0).^42^ This has been used previously in comparable biomedical settings.^43,44^ Feature expansion was only applied to features that had missing values. There were no missing values for ICD and OPS codes, systemic treatments, cancer diagnoses, age, and sex.

### External Flatiron Health dataset

This study used the nationwide Flatiron Health electronic health record (EHR)-derived de-identified database. The Flatiron Health database is a longitudinal database, comprising de-identified patient-level structured and unstructured data, curated via technology-enabled abstraction.^45,46^ During the study period, the de-identified data originated from approximately 280 cancer clinics (∼800 sites of care). The study included 3,288 patients diagnosed with advanced non-small cell lung cancer (NSCLC) from 01 January 2011 to 10 November 2022. The majority of patients (82.7%) originate from community oncology settings. The data are de-identified and subject to obligations to prevent re-identification and protect patient confidentiality. Patients with a birth year of 1937 or earlier may have an adjusted birth year in Flatiron datasets due to patient de-identification requirements.

For subsequent analysis in this study, extreme outliers were discarded manually before outliers, defined as >3 standard deviations from the mean, were removed for continuous parameters. Further preprocessing of the data was performed analogously to the internal dataset. This resulted in a total of 18 parameters for the final validation dataset.

### Model architecture

To model treatment outcomes, we used the coxph architecture similar to DeepSurv and the training regime from the pycox survival library.^5,47^

Each parameter (potentially feature-expanded) was used as an input to a fully connected neural network with one hidden layer and a hidden width of 10 times the input neurons.

Thus, we decided to follow an early-fusion approach since (1) all markers are 1-dimensional and reasonably independent from each other (unlike, for example, pixels of an image or DNA sequences used in other studies) and (2) early fusion is particularly suitable for allowing interactions between markers.^48^

### Model Training

Using five-fold cross-validation, we trained, for each fold, two neural networks (OS, TTNT) on 80% of the data to predict the proportional hazard risk score for the overall survival (OS) and time to next treatment (TTNT), respectively. We used the training algorithm supplied by the pycox library.^47^ The remaining 20% of data was split randomly into a validation set (10%) to early-stop the model and a test set (10%) for the computation of the concordance index. Cancer entities were balanced between training and validation/test sets for each fold.

Models were trained for up to 50 epochs with a learning rate of 0.01 using the Adam optimizer. We used the default early stopping algorithm supplied by pycox. After the training process was early stopped, the learning rate was reduced to 1/10 of the previous learning rate and the model was trained for another 50 epochs. This was repeated down to a learning rate of 1e-4. We used a dropout rate of 0.5 and a batch size of 1024. To reduce the effect of correlations between input parameters on the relevance explanation, we applied input dropout at a rate of0.5 during training.^40^ The concordance scores between predicted risk and ground truth were calculated for each fold using the pycox library. The identical training, validation, and test splits were used when neural networks were trained on individual cancer entities compared to training on the pan-cancer dataset to ensure comparability. Concordance results were discarded if the test set consisted of less than ten samples or if the test samples did not have at least five events.

### Explaining ML Predictions

To explain the model’s predictions, we used layer-wise relevance propagation (LRP), a method for xAI that leverages the neural network structure of the model to compute explanations robustly and efficiently.^12^ LRP starts with the prediction (the value obtained at the output of the neural network) redistributes it backwards, layer after layer, by means of propagation rules, and collects the explanation in the input layer. A physical analogy to the LRP propagation is water flowing through a network of pipes. In this physical network, the amount of water injected at the output equals the amount observed at the input.

More formally, let j and k be indices for neurons in two consecutive layers and *a*_*j*_ and *a*_*k*_ be their respective activations. In a typical neural network, including the DeepSurv network considered in this work, two consecutive layers are related generically by the equation:

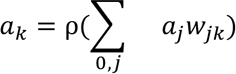

In this equation, the sum runs over all neurons in the given layer plus a neuron with constant activation *a*_0_ = 1. The variable *W*_*jk*_ is the weight connecting neuron *j* to neuron *k*. We then backpropagate using the generalized LRP-gamma rule, similar to previous works.^43,44^ This rule propagates from one layer to the layer below using the equation:

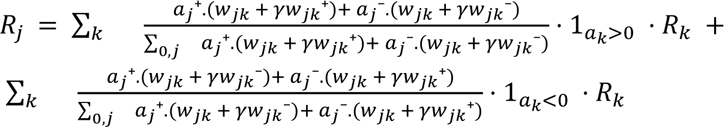

where (.)^+^ = *max*(0, .) and (.)^−^ = *min*(0, .), and where *γ* is a parameter that needs to be selected. Here, we used the heuristic 0.01 which worked well in other applications.^44^ Applying the rule at each layer, starting at the top layer and moving backwards until the input layer, we obtain in the last step the contribution of each input feature to the prediction. For expanded features, the final LRP score is calculated as the sum of the LRP scores assigned to the tuple (x, 1-x).

We treated the LRP score assigned to a specific input as the risk contribution (RC) of this marker to the overall patient prognosis (OS or TTNT). The ‘marker importance’ of a marker across all patients was defined as the sum of the absolute LRP scores divided by the number of patients for whom this marker was not missing. To calculate the marker importance in a subcohort (e.g., patients of a single cancer entity), LRP scores were first centered by subtracting the cohort mean.

### Statistics

The statistical analyses were conducted in R statistical packages.^49^ All tests were two-sided and results were regarded as significant if p<0.05. Model performance (**Fig. 2**) was compared using Wilcoxon ranked test in the package Hmisc.^50^

Linear regression was applied to fit relationships between marker values and their corresponding xAI-assigned RC for the internal and external datasets, respectively. Subsequently, the slope coefficients of these models were compared between the internal and external datasets.

The search for interactions between markers was quantified by comparing linear mixed-effects models with baseline models. For each marker pair, the relationship between the ‘primary’ marker and the RC was examined under the two conditions when the ‘secondary’ marker was high (highest 10%) or low (lowest 10%). For categorical parameters, category levels were selected so that at least 10% of the samples were members of the high or low class, respectively. Medications, ICD codes, OPS codes, and cancer types were excluded from this analysis due to unbalanced levels. Marker pairs that were present in less than 100 samples were discarded. Holm’s multiple test correction was applied.

To validate marker relationships of higher complexity, we examined marker pairs that were found in the internal and external datasets. The difference in model coefficients between ‘high’ and ‘low’ classes was compared between both datasets. This analysis was restricted to markers that were present in both datasets. For the simple linear model, the baseline was a model consisting of the intercept only. For the mixed effects linear model, the baseline consisted of a linear model with a fixed slope and a random intercept.

Additionally, these relationships between marker values and RC were compared with the coefficients (i.e., hazard ratios) of univariate Cox proportional hazard models that predicted survival based on the respective markers. A mixed-effects variant of Cox proportional hazards models was used to validate the mixed-effects case. Cox models were discarded if they had a lower log-likelihood than their baseline models, but did not have to be significant to be included in the comparison.

Cox proportional hazards models were implemented with the Survival package.^51^ The mixed-effects variants of this analysis were modeled using the coxme package.^52^ Other mixed-effects models were implemented with lme4.^53^

## Figures

Kaplan-Meier plots were computed with the R package *survival*.^54^ Fig. 1 was created with BioRender.com. All other plots were created with *ggplot2*.^55^ For Fig. 2, only cancer ICD codes were selected for which there were at least 20 patients in each fold’s test set. For cancers with fewer patients, the training on only the specific cancer ICD code (for comparison against training on all cancer entities) became unstable. For Fig. 2B, patients from the test sets were stratified based on the models trained and validated on training and validation sets. This was repeated for each fold.

Fig. 3 shows marker values and corresponding RC for all patients.

Fig. 4 shows a selection of features for selected patients.

Fig. 5 **and Suppl.** Fig. 11 only show cancer entities for which the respective marker has been measured in at least 20 patients.

Fig. 6 **and Suppl Fig. 12-14**: Basis of the xKM plot is a conventional Kaplan-Meier plot, including all patients who were not censored. For each marker, the marker importance curve was added by calculating a regression line through the marker importance score for each event. The xKM plots were limited to a 2-year time window for better visualization. To make the results more robust, only markers measured in at least 40 patients are shown.

**Suppl.** Fig. 2 shows the coefficients of the linear models between marker values and RC and compares them between the internal and external datasets.

**Suppl.** Fig. 3 compares the coefficients from **Suppl.** Fig. 2 to the coefficients fitted in the Cox proportional hazards scores.

**Suppl.** Fig. 4 shows marker values and corresponding RC for all patients.

**Suppl.** Fig. 5 is similar to **Suppl.** Fig. 2, but instead of showing linear model coefficients, it shows the difference in random effect coefficients between high and low ‘secondary markers’. The labels X->Y indicate the difference in the influence of marker Y on the prognosis when the ‘secondary marker’ X is high versus low.

**Suppl.** Fig. 6 is similar to **Suppl.** Fig. 3, but instead of showing linear model coefficients, it shows the differences in random effect coefficients between high and low ‘secondary markers’. The labels X->Y indicate the difference in the influence of marker Y on the prognosis when the ‘secondary marker’ X is high versus low.

**Suppl.** Fig. 7 only shows cancer entities for which each marker was measured in at least 20 patients. In addition, a marker value (e.g., ECOG score of 3) was discarded if no other patient in the same cancer entity had the same value for that marker.

**Suppl.** Fig. 8 shows the importance of each marker across all patients. The markers had to be measured in at least 20 % of the cancer entities in at least 10 % of the patients. Otherwise, they were presented in a transparent font.

**Suppl.** Fig. 9 shows the importance of the 70 most important markers across all patients. Rare and particularly tumor-specific markers were discarded in this overview according to the following threshold: Markers had to be measured in at least 20% of cancer entities in at least 10% of patients. This analysis does not include information about ICD and OPS codes or systemic treatments.

**Suppl.** Fig. 10 shows the RC of ICD and OPS codes. Markers are ordered from top to bottom by decreasing mean RC and only the first 70 markers are shown.

## Acknowledgements

J.Keyl is grateful for the support of the Clinician Scientist Academy of the University Hospital Essen (UMEA).

## Funding

This work was partly funded by the German Ministry for Education and Research (under refs 01IS14013A-E, 01GQ1115, 01GQ0850, 01IS18056A, 01IS18025A and 01IS18037A) and BBDC/BZML and BIFOLD. Furthermore, K.-R.M. was partly supported by the Institute of Information & Communications Technology Planning & Evaluation (IITP) grants funded by the Korea Government (MSIT) (No. 2019-0-00079, Artificial Intelligence Graduate School Program, Korea University and No. 2022-0-00984, Development of Artificial Intelligence Technology for Personalized Plug-and-Play Explanation and Verification of Explanation).

J.H. received financial support by the German Research Foundation (DFG) funded Clinician Scientist Academy of the University Hospital Essen (UMEA) (FU 356/12-2).

A.R. was in part funded by the Deutsche Forschungsgemeinschaft (DFG, German Research Foundation) – Project-ID 418179183 – KFO 337(RO 3577/3-2 (AR), RO 3577/7-1 (AR), SCHA 422/17-1 (DS).

JTS is grateful for support by the German Cancer Consortium (DKTK) and by the German Federal Ministry of Education and Research (BMBF; 01KD2206A/SATURN3).

## Contributions

Conceptualization: J.Keyl and P.K.

Methodology: J.Keyl, P.K., G.M.

Formal analysis: J.Keyl and P.K. Investigation: all authors.

Resources: M.Schuler., S.B., N.B., M.F., D.F.-S., M.G., V.G., B.H., J.H., K.H., S.K., R.K., S.L.,

T.R., A.R., D.S., J.T.S., M.Stuschke, U.S., M.T., A.W., M.W., H.A.B., F.N.

Data curation: J.Keyl, R.H. Writing—original draft: J.Keyl and P.K.

Writing—review and editing: all authors.

Visualization: J.Keyl, P.K., A.B.

Supervision: J.Kleesiek, F.K., M.Schuler and K.-R.M.

## Conflict of interest statement

**V.G.:** Honoraria: Bristol Myers Squibb, Pfizer, Ipsen, Eisai, Merck Sharp & Dohme (MSD) Oncology, Merck HealthCare, EUSAPharm, Apogepha, Ono Pharmaceutical. Advisory role: BMS, Pfizer, MSD Oncology, Merck HealthCare, Ipsen, Eisai, Debiopharm, PCI Biotech, Cureteq, Oncorena. Travel: Pfizer, Ipsen, Merck HealthCare.

**B.H.:** Advisory role: ABX, AAA/Novartis, Astellas, AstraZeneca, Bayer, BMS, Janssen R&D, Lightpoint Medical, Inc., and Pfizer. Research funding: Astellas, BMS, AAA/Novartis, German Research Foundation, Janssen R&D, and Pfizer. Travel: Astellas, AstraZeneca, Bayer, and Janssen.

**D.S.:** Personal fees for advisory boards: BMS, Immunocore, MSD, Neracare, Novartis, Pfizer, Philogen, Pierre Fabre, Sanofi/Regeneron. Personal fees as an invited speaker: BMS, Merck Serono, MSD, Novartis, Roche, Sanofi. Personal fees (financial interest) for steering committee membership: BMS, MSD. Personal support (no financial interest) for steering committee membership: Novartis. Institutional support as a coordinating principal investigator (no financial interest): BMS, MSD, Novartis, Pierre Fabre. Institutional support as a local principal investigator (no financial interest): Philogen, Sanofi. Institutional research grant support (financial interest): BMS, MSD. EORTC-MG Member of Board of Directors (no financial interest).

**J.T.S.:** Receives honoraria as consultant or for continuing medical education presentations from AstraZeneca, Bayer, Boehringer Ingelheim, Bristol-Myers Squibb, Immunocore, MSD Sharp Dohme, Novartis, Roche/Genentech, and Servier. His institution receives research funding from Abalos Therapeutics, Boehringer Ingelheim, Bristol-Myers Squibb, Celgene, Eisbach Bio, and Roche/Genentech; he holds ownership and serves on the Board of Directors of Pharma15, all outside the submitted work.

**M.T.:** Speaker fees and personal support: AstraZeneca, Daiichi Sankyo, Novartis, Bayer, Asklepios, Edwards LifeSciences.

**M.W.:** Honoraria and advisory role: Amgen, AstraZeneca, Daiichi Sankyo, GlaxoSmithKline, Janssen, Novartis, Pfizer, Roche, Takeda. Research funding: Bristol-Myers Squibb, Takeda.

**M.S.:** Consultant (compensated): Amgen, AstraZeneca, Blueprint Medicines, Boehringer Ingelheim, Bristol-Myers Squibb, GlaxoSmithKline, Janssen, Merck Serono, Novartis, Roche, Sanofi, Takeda. Stock ownership: None. Honoraries for CME presentations: Amgen, Boehringer Ingelheim, Bristol Myers Squibb, Janssen, MSD, Novartis, Roche, Sanofi. Research funding to institution: AstraZeneca, Bristol-Myers Squibb.

## Data Availability Statement

Anonymized data are available upon reasonable request. Data cannot be shared with investigators outside the institution without consent. The data that support the findings of this study have been originated by Flatiron Health, Inc. Requests for data sharing by license or by permission for the specific purpose of replicating results in this manuscript can be submitted to.

## Notes

### Funding Statement

J.Keyl is grateful for the support of the Clinician Scientist Academy of the University Hospital Essen (UMEA).
This work was partly funded by the German Ministry for Education and Research (under refs 01IS14013A-E, 01GQ1115, 01GQ0850, 01IS18056A, 01IS18025A and 01IS18037A) and BBDC/BZML and BIFOLD. Furthermore, K.-R.M. was partly supported by the Institute of Information & Communications Technology Planning & Evaluation (IITP) grants funded by the Korea Government (MSIT) (No. 2019-0-00079, Artificial Intelligence Graduate School Program, Korea University and No. 2022-0-00984, Development of Artificial Intelligence Technology for Personalized Plug-and-Play Explanation and Verification of Explanation).
J.H. received financial support by the German Research Foundation (DFG) funded Clinician Scientist Academy of the University Hospital Essen (UMEA) (FU 356/12-2).
A.R. was in part funded by the Deutsche Forschungsgemeinschaft (DFG, German Research Foundation) - Project-ID 418179183 - KFO 337(RO 3577/3-2 (AR), RO 3577/7-1 (AR), SCHA 422/17-1 (DS).
JTS is grateful for support by the German Cancer Consortium (DKTK) and by the German Federal Ministry of Education and Research (BMBF; 01KD2206A/SATURN3).

### Author Declarations

The study was approved by the Ethics Committee of the Medical Faculty of the University of Duisburg-Essen (No. 21-10347-BO).

### Summary of Updates

Updated Metadata details; Supplementary files updated; Format changes

