## Supplementary Material for "Decoding pan-cancer treatment outcomes using multimodal real-world data and explainable artificial intelligence"

**Supplementary Figure 1**: Flowchart showing the process of patient enrollment.

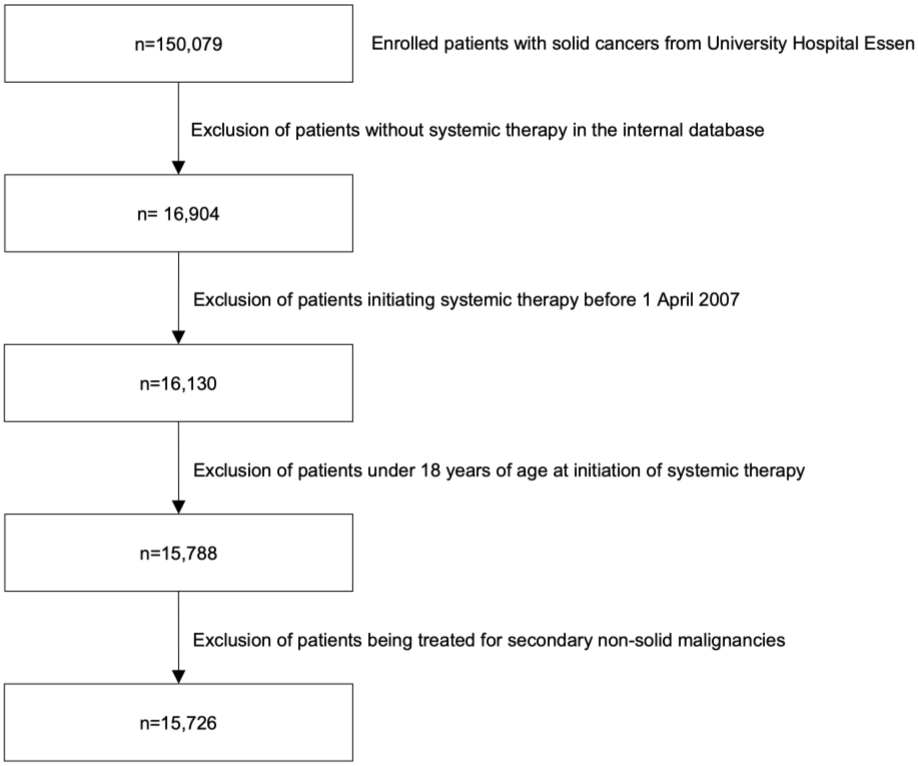


**Supplementary Table 1**: Composition of the patient cohort.

| Cancer | ICD | Patients |
|---|---|---|
| Lung | C34 | 4320 |
| Sarcoma | C40, C41, C47-C49 | 1578 |
| Breast | C50 | 1223 |
| Head and Neck | C01-C13, C30-C32 | 1026 |
| Liver | C22 | 728 |
| Brain | C71 | 644 |
| Pancreas | C25 | 620 |
| Colon | C18 | 606 |
| Melanoma | C43 | 600 |
| Stomach | C16 | 545 |

| | | |
|---|---|---|
| Esophagus | C15 | 408 |
| Eye | C69 | 384 |
| Rectum | C20 | 373 |
| Kidney | C64, C65 | 308 |
| Uterus | C53-C55 | 275 |
| Testis | C62 | 249 |
| Prostate | C61 | 236 |
| Mesothelioma | C45 | 229 |
| Skin | C44 | 217 |
| Bladder | C67 | 166 |
| Biliary tract | C24 | 163 |
| Ovary | C56 | 152 |
| Thyroid gland | C73 | 117 |
| Small intestine | C17 | 90 |
| Heart | C38 | 84 |
| Rectosigmoid junction | C19 | 80 |
| Gallbladder | C23 | 64 |
| Anus | C21 | 48 |
| Other endocrine gland | C75 | 34 |
| Urethra | C68 | 33 |
| Vulva | C51 | 29 |
| Other digestive organs | C26 | 24 |
| Thymus | C37 | 21 |
| Adrenal gland | C74 | 17 |
| Penis | C60 | 11 |
| Ureter | C66 | 10 |
| Female genital | C57 | 9 |
| Central nervous system | C72 | 5 |

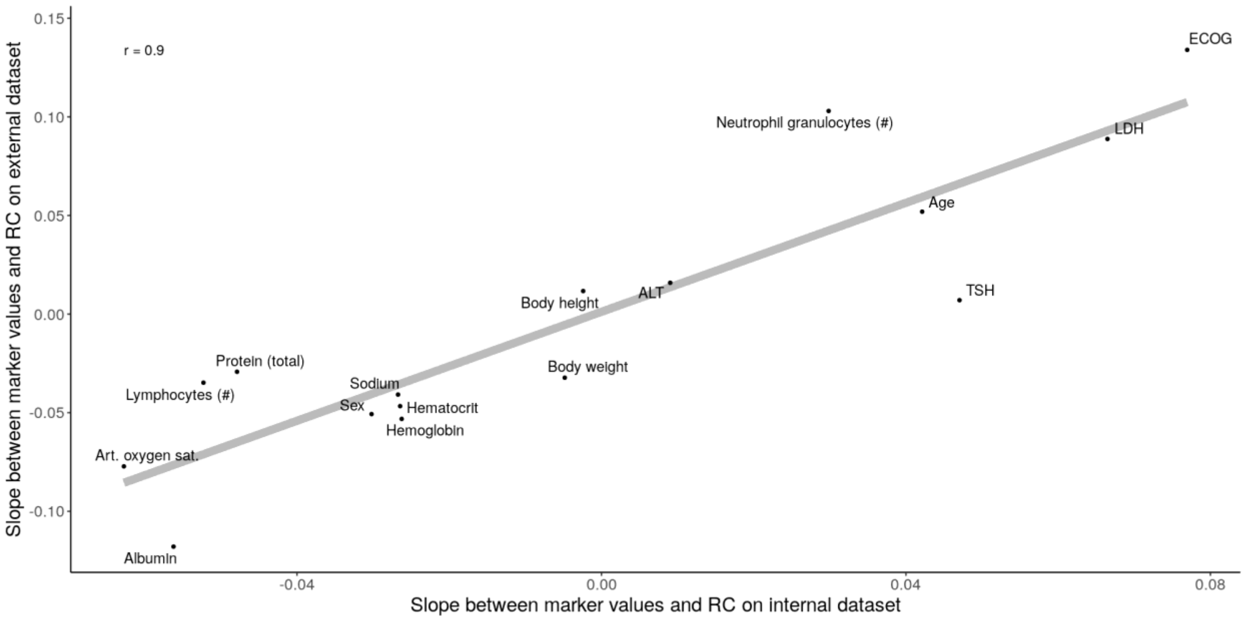


**Supplementary Figure 2:** Replicability of the xAI approach on the external dataset. Axes indicate the (linearized) relationship between marker values and their xAI-assigned RCs for the Internal (x axis) and external (y axis) datatset. xAI finds the similar relationships in both datasets (r=0.9).

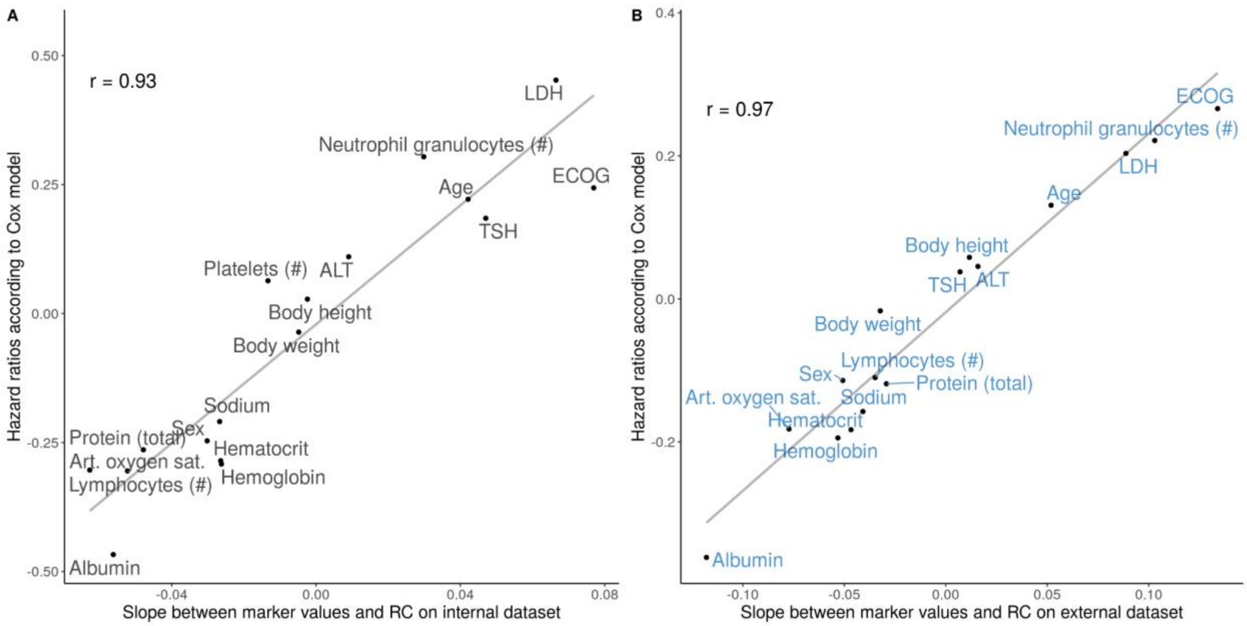


**Supplementary Figure 3:** Validation of xAI results with Cox regression models. The x axis shows the linearized relationships between marker values and RC according to xAI (similar to Supplementary Figure 2). The y axis shows the hazards of each marker according to a univariate cox regression model on the same dataset. The cox regression models strongly support the results captured by xAI. A: Internal dataset (Pearson's r = 0.93), B: External dataset (Pearson's r = 0.97).

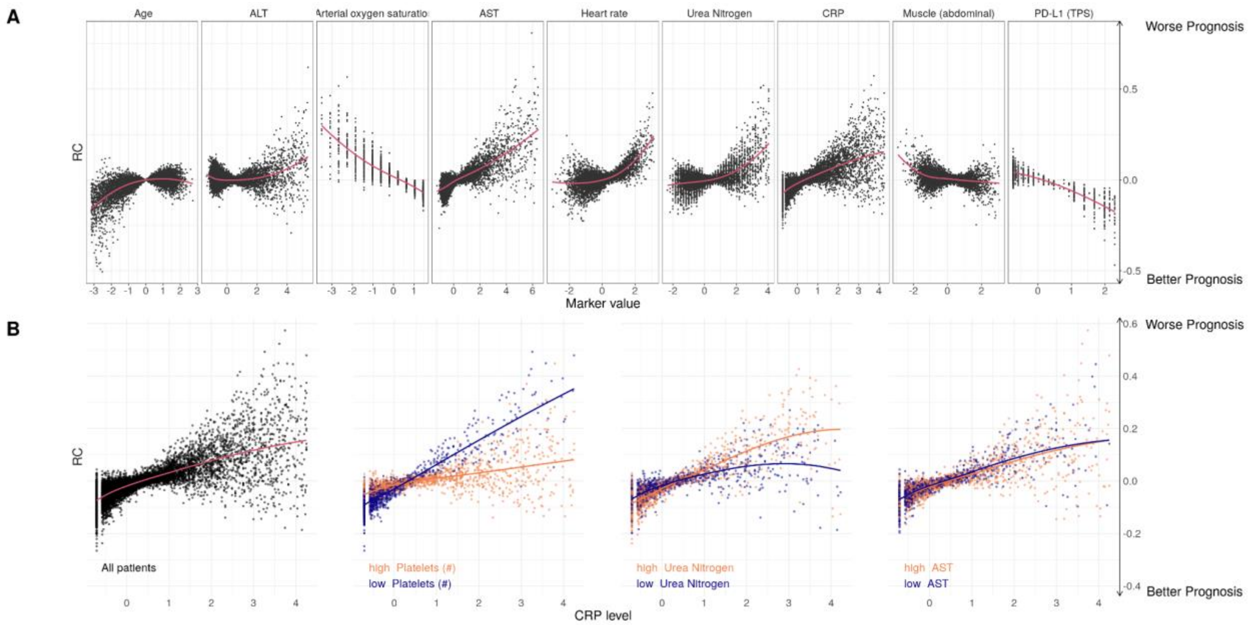


**Supplementary Figure 4:** Contribution of clinical markers to the prediction of TTNT.

**A:** Marker risk contribution (RC) on the TTNT prediction. Each point represents one marker value for one patient versus the LRP-assigned RC (y-axis) to the patient's prognosis. Marker values are standardized.

**B:** The risk contribution of CRP depended on the value of other markers. The standardized CRP level and LRP-assigned RC are shown for all patients in the left plot. Right three plots depict the patients for whom the three selected markers platelet count, urea nitrogen and AST were in the highest or lowest 10% quantile.

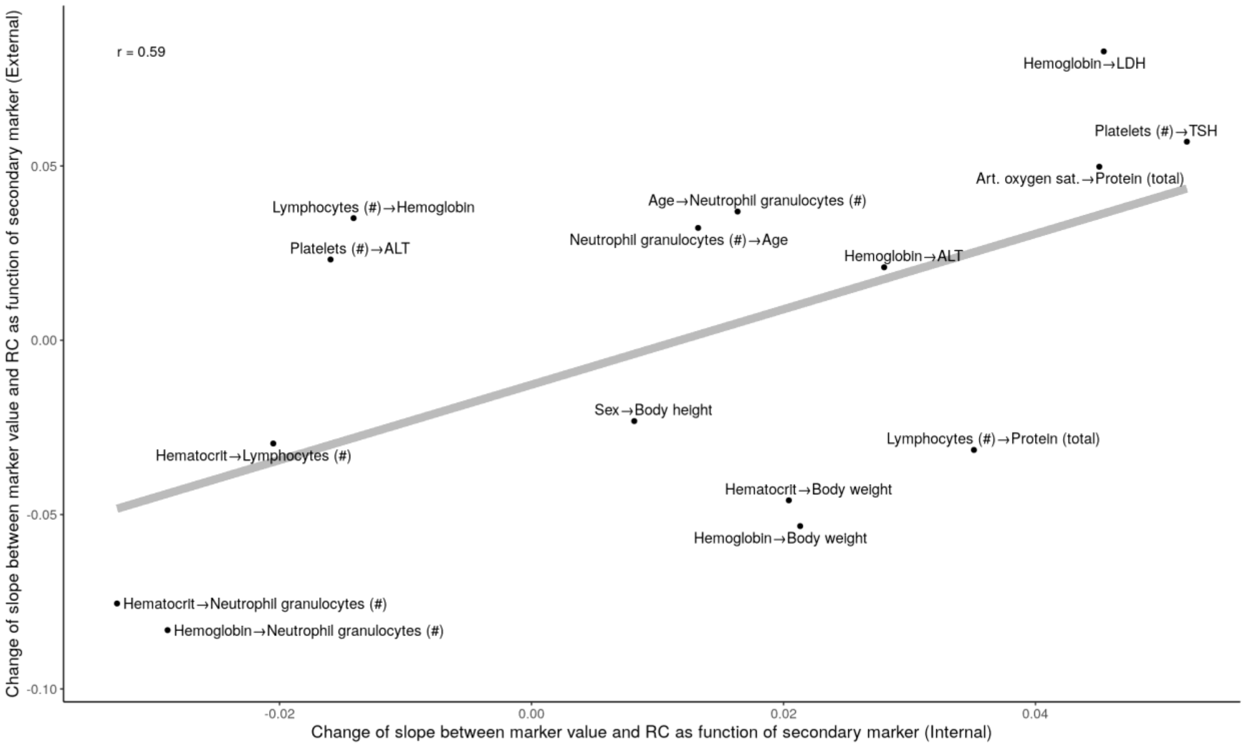


**Supplementary Figure 5:** Axes indicate how the (linearized) relationships between marker values and xAI-assigned RCs differ between patient subgroups that depend on other markers (high or low). Given the linearized relationship between a marker Y and the RC of Y, the label X->Y defines how this relationship changes between patient groups with high and low X. Even these higher order interactions between markers show similar patterns in the internal (x axis) and external (y axis) dataset (r=0.58).

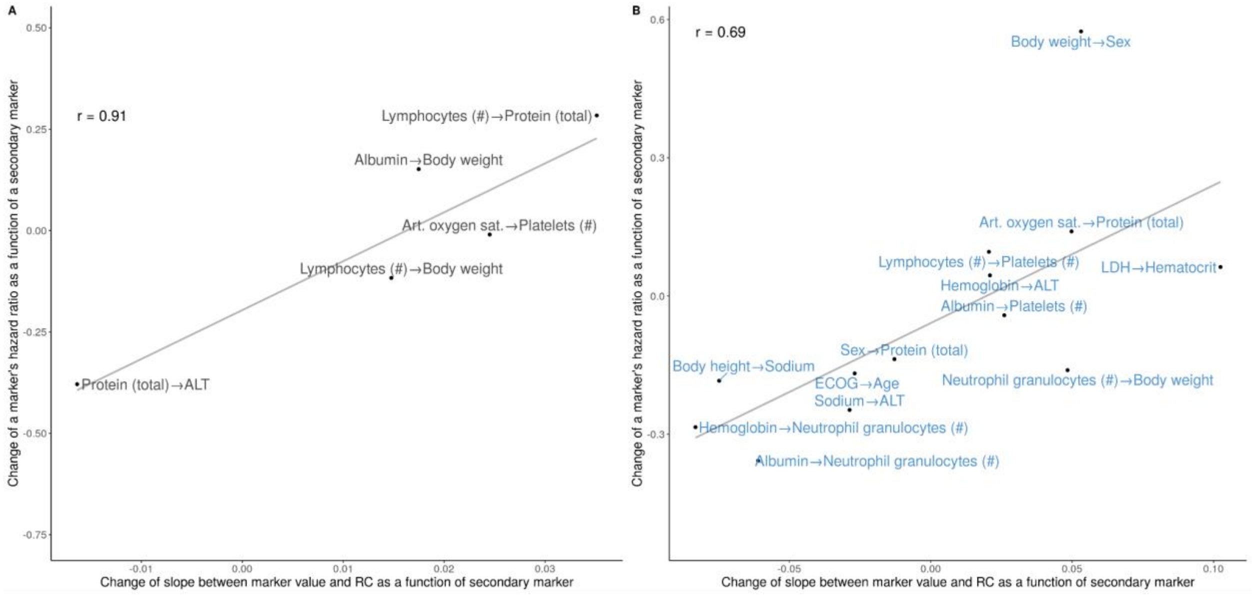


**Supplementary Figure 6:** Complex interactions found by xAI can be validated with mixed-effects Cox proportional hazards models. Given the linearized relationship between a marker Y and the RC of Y, the label X->Y defines how this relationship changes between patient groups with high and low X. The effects captured by xAI (x axis) correspond strongly to the effects estimated by mixed-effects Cox proportional hazards models. **A**: Internal dataset (r=0.91). **B**: External dataset (r=0.69).

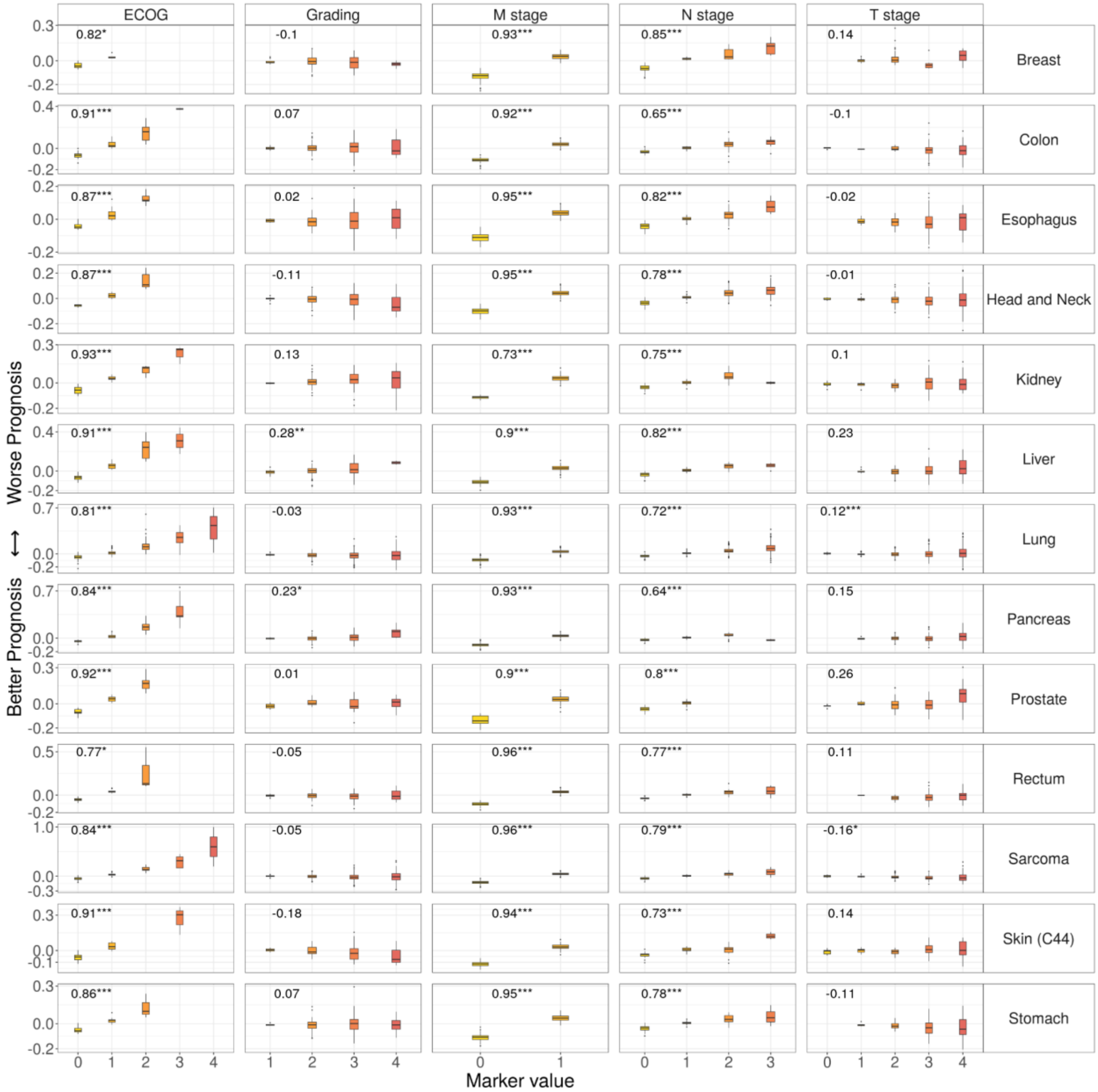


**Supplementary Figure 7:** Comparison of established prognostic scores with the LRP-assigned RC for OS. The x-axis depicts the value of the different scores. The y-axis indicates the RC. Comparison is shown for each marker and cancer type. Asterisks indicate if correlation was significant. *: p<0.01, **: p<0.001, ***: p<0.0001

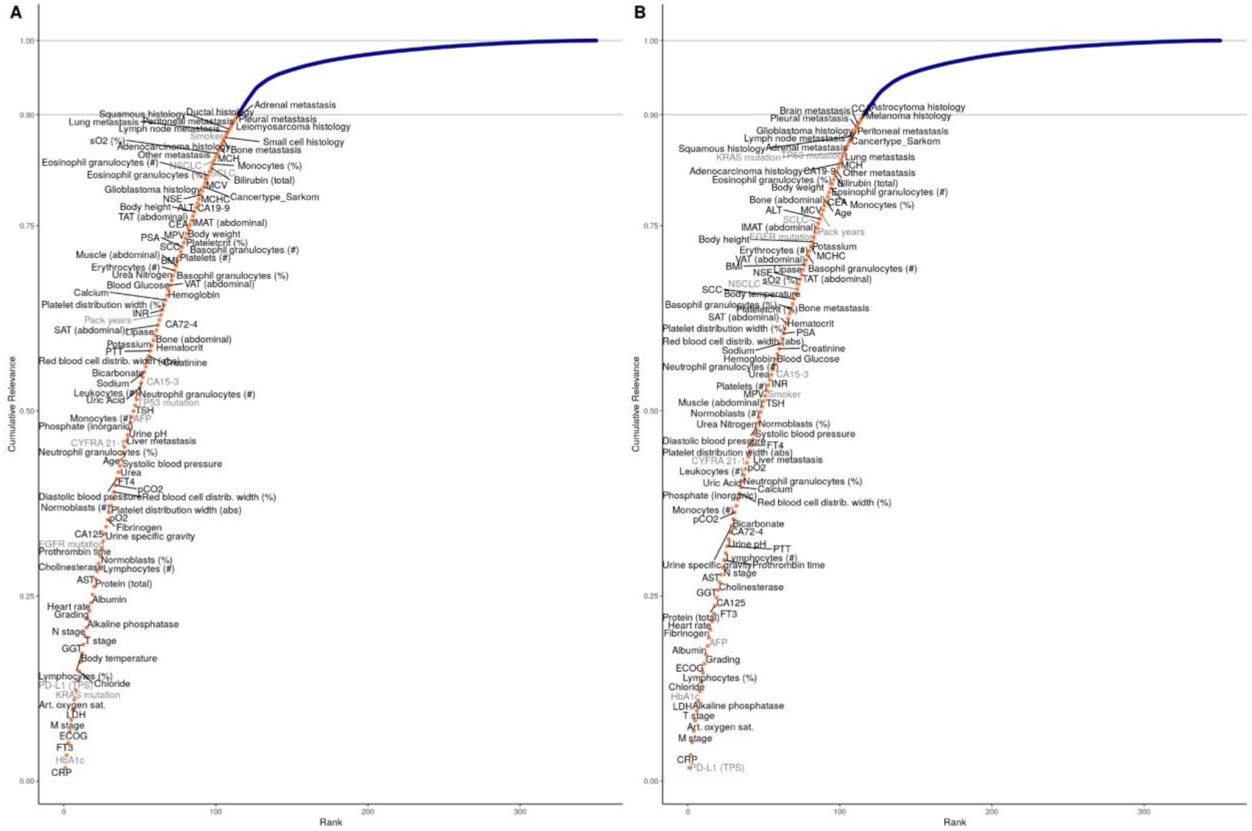


**Supplementary Figure 8**: Cumulative relevance for neural network decision-making. A: OS, B: TTNT
All markers are ranked according to the decreasing marker importance (MI) assigned by LRP across all patients (x axis). MI is corrected for missing values. Y-axis shows the cumulative MI. 90 % of all MI is assigned to 114 (TTNT: 115) key prognostic markers.

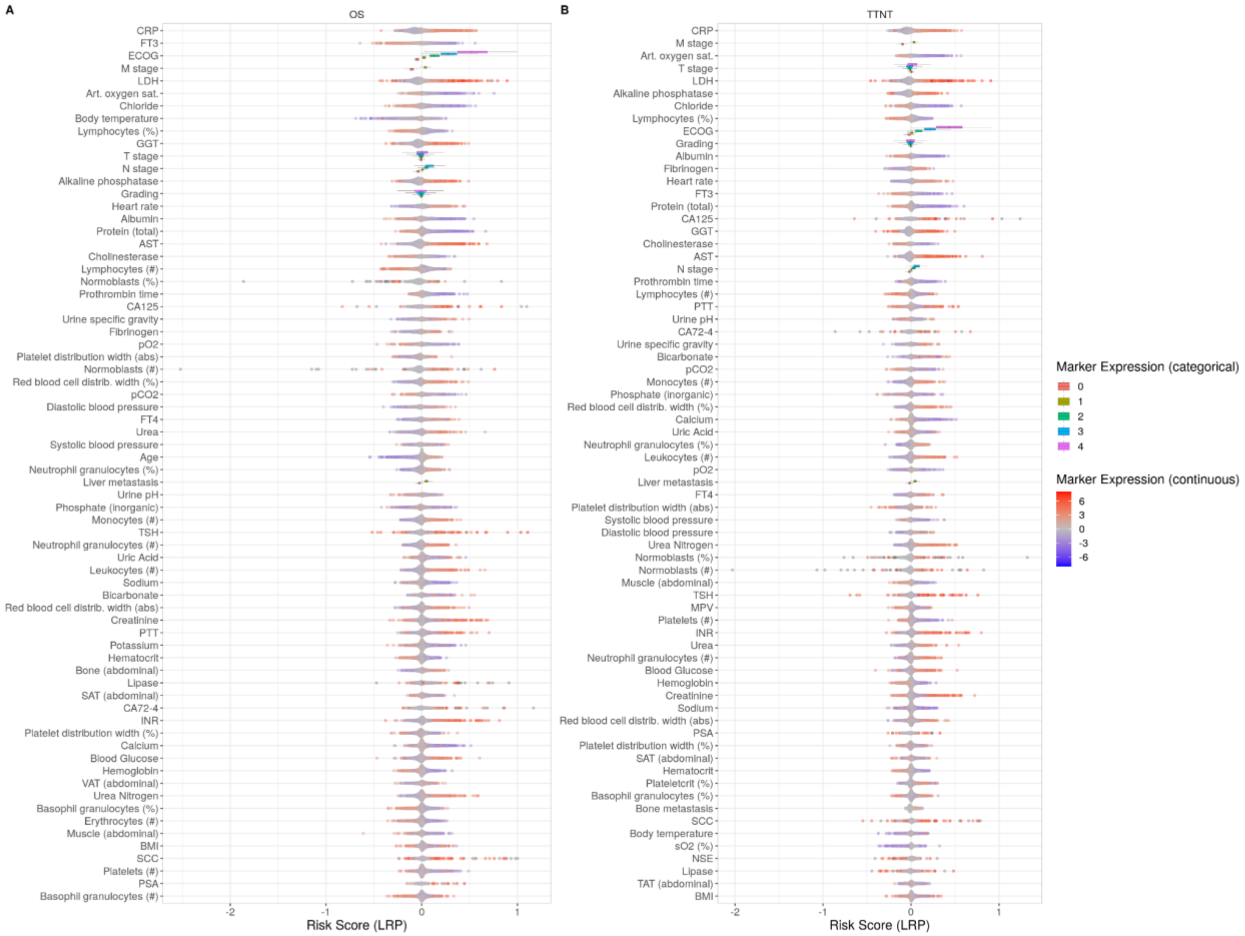


**Supplementary Figure 9**: Marker importance of diagnostic features. Markers are ordered from top to bottom according to decreasing importance across all patients. Points are measurements in individual patients. Risk contribution (RC) of markers in individual patients is shown on the x axis. RC indicates the contribution to a better (negative) or worse (positive) prognosis. Point color indicates high (red) or low (blue) marker value. **A:** Contribution to overall survival (OS). **B:** Contribution to time-to-next-treatment (TTNT).

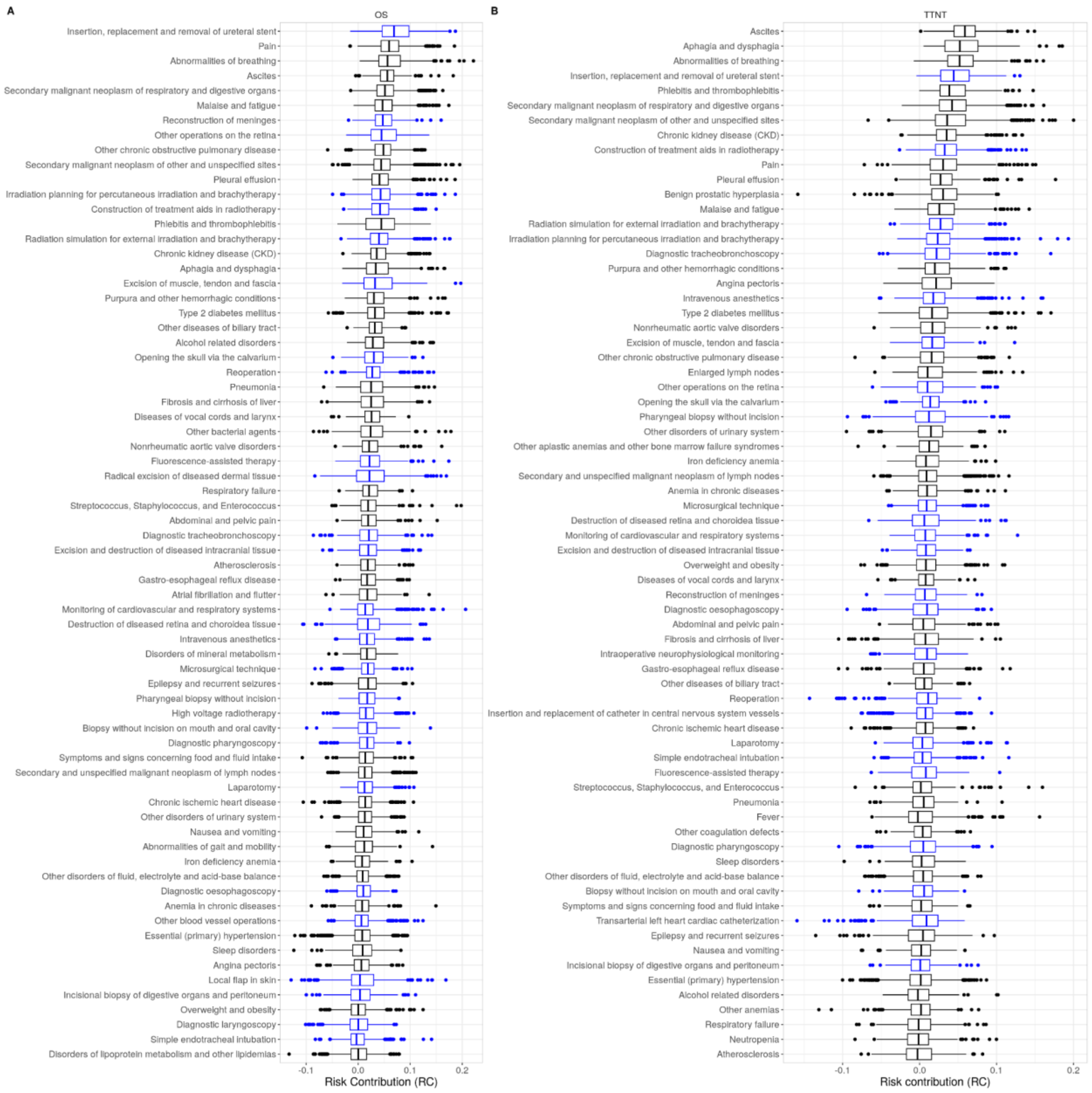


**Supplementary Figure 10**: ICD (black) and OPS codes (blue) with the highest assigned RC. **A:** Contribution to overall survival (OS). **B:** Contribution to time-to-next-treatment (TTNT).

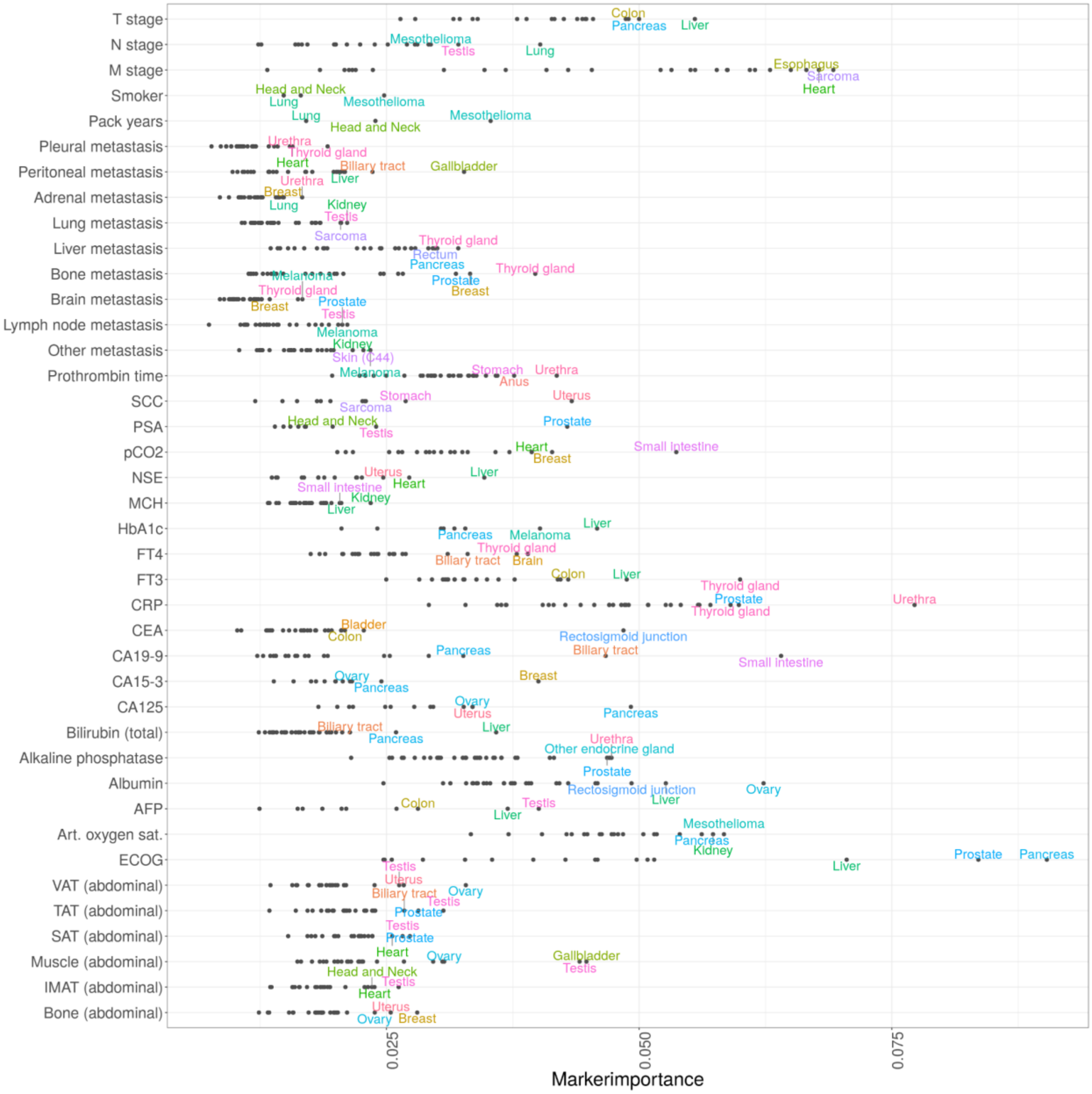


**Supplementary Figure 11:** The relationship between mean marker importance (MI) of selected markers and cancer entities for TTNT. The x axis shows the MI on a logarithmic scale. For each marker, the three cancer entities with the highest marker MI are annotated. Body composition markers: Abdominal volumes of visceral adipose tissue (VAT), total adipose tissue (TAT), subcutaneous adipose tissue (SAT), intermuscular adipose tissue (IMAT), muscle, bone. ECOG PS had the highest MI in pancreatic, prostate, and liver cancers, whereas fT3 was particularly important for thyroid, testicular, and brain cancers.

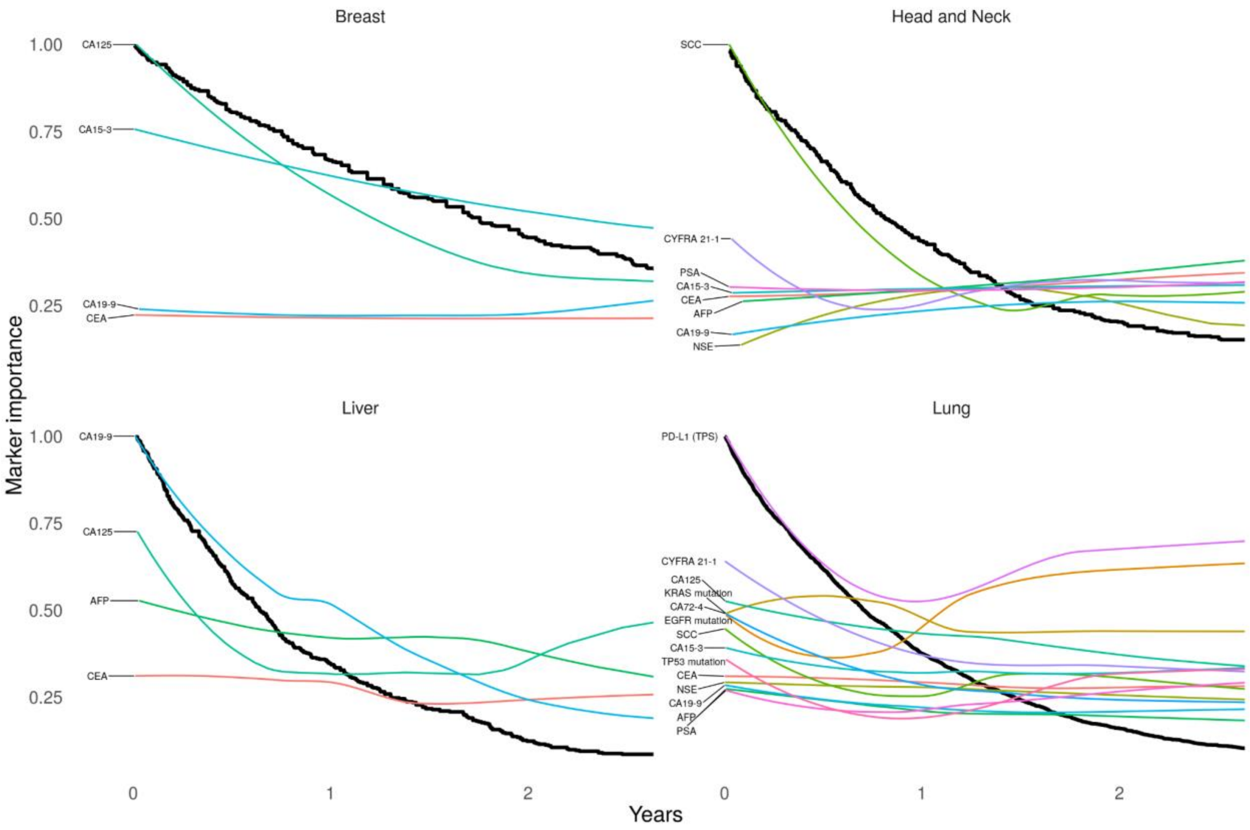


**Supplementary Figure 12:** xKM curves show the progress of marker contribution for the prediction of overall survival (OS) for tumor-specific markers along disease progression. Black lines represent Kaplan-Meier plots, while the colored lines visualize the change in marker importance (MI) for patients with different survival times. MI lines are scaled between zero and one. Only deceased patients were included in this analysis.

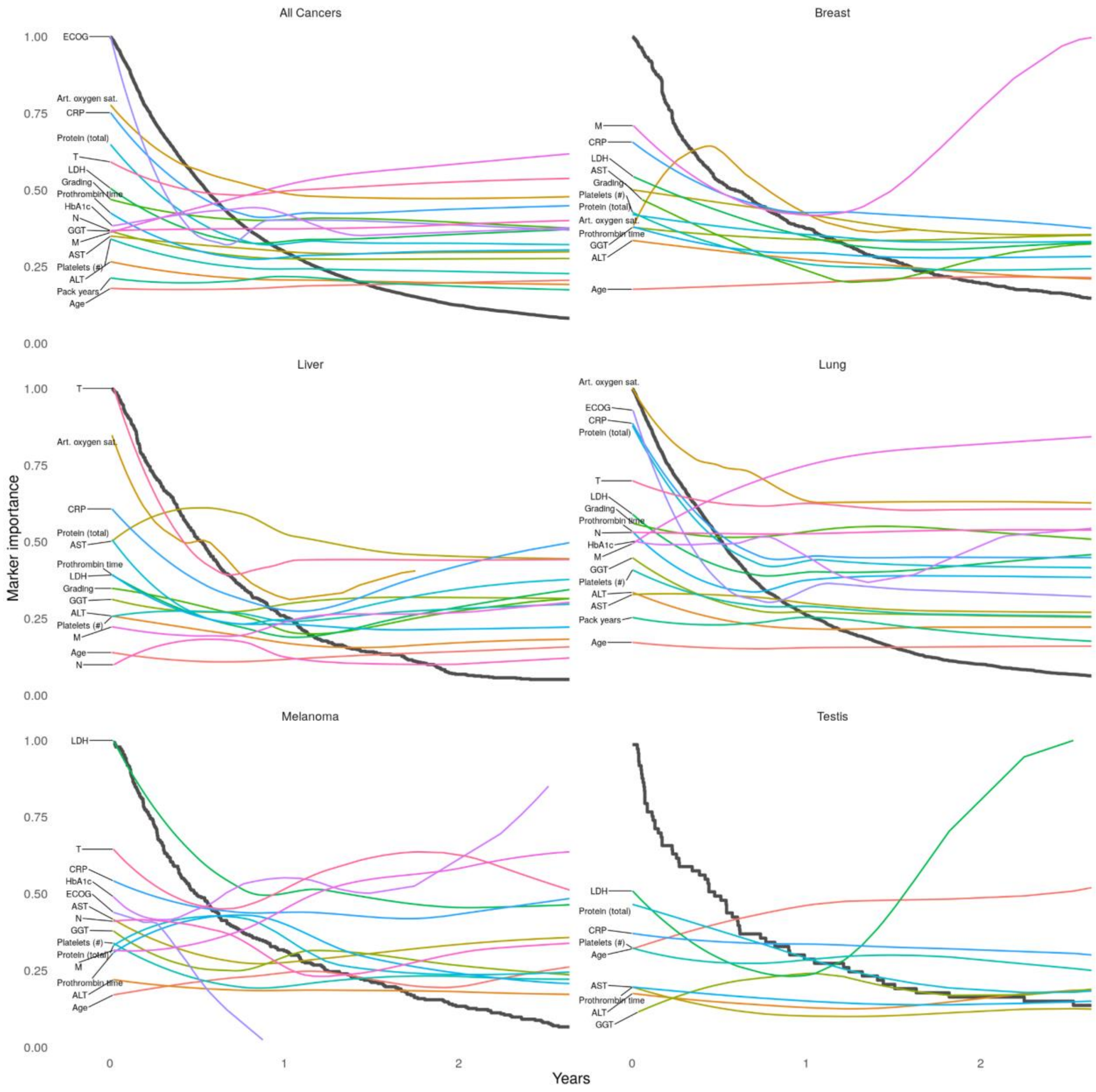


**Supplementary Figure 13:** Explainable Kaplan-Meier (xKM) plots depicting the importance of diagnostic markers for TTNT during disease progression. Black lines represent Kaplan-Meier plots, while the colored lines visualize the change in marker importance (MI) for patients with different survival times. MI lines are scaled between zero and one. Only deceased patients were included in this analysis.

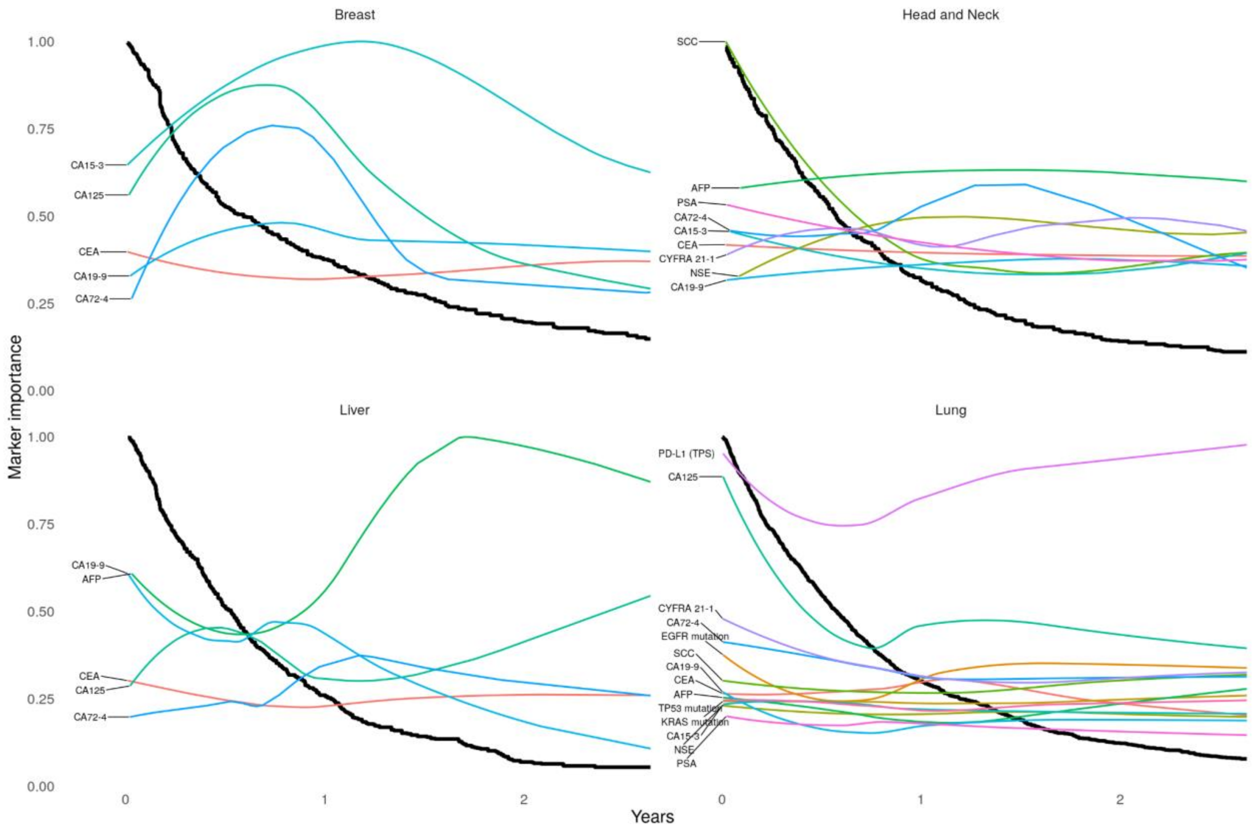


**Supplementary Figure 14:** xKM curves show the progress of marker contribution for the prediction of time-to-next-treatment (TTNT) for tumor-specific markers along disease progression. Explainable Kaplan-Meier (xKM) plots depicting the importance of diagnostic markers during disease progression. Black lines represent Kaplan-Meier plots, while the colored lines visualize the change in marker importance (MI) for patients with different survival times. MI lines are scaled between zero and one. Only deceased patients were included in this analysis.
